# Reliability of 3T quantitative human brain MRI is preserved after scanner relocation

**DOI:** 10.64898/2026.09.17.26363342

**Authors:** Stefano Tambalo, Alberto Finora, Paula A. Maldonado Moscoso, Manuela Moretto, Sebastian Hübner, Lara Maria Viola, Manuela Piazza, Nicola Pace Tom Hilbert, Gian Franco Piredda, Tobias Kober, Jorge Jovicich

## Abstract

Quantitative magnetic resonance imaging (qMRI) supports both basic neuroscience and clinical research, providing the foundation for longitudinal studies of brain development, aging, and neurological disease. However, its validity rests on a little-studied assumption: that physical relocation of imaging hardware leaves underlying neuroimaging biomarkers intact. When research facilities expand or modernize, an entire scanner undergoes radical disruption—field ramp-down, physical transport, shielding reconstruction, and re-shimming. Whether qMRI metrics remain stable following such invasive procedures without introducing systematic biases is largely unquantified. Here, we systematically evaluated the impact of relocating a 3T clinical scanner over a distance of 30 km on a broad, multimodal suite of neuroimaging markers in healthy volunteers. To disentangle variance components, we benchmarked within- and cross-site stability using percent variation (PV; measurement precision), intraclass correlation coefficients (ICC; absolute reliability), and Spearman rank correlations (R^2^; spatial and order agreement). We show that scanner relocation introduces no greater variance than baseline within-site assessments. High-resolution brain structural morphometry from different sequences (multi-echo MPRAGE, compressed sensing MPR2RAGE) and quantitative relaxometry (compressed sensing MP2RAGE T1 mapping) demonstrated excellent stability (PV<1%, ICC=0.89-0.99, R^2^>0.93). Diffusion tensor (mean diffusivity: MD, fractional anisotropy: FA) and mean signal kurtosis (MSK) metrics from multi-shell diffusion MRI maintained low variability across white and deep gray matter (global cross-site ICC>0.90; global PV: MD<1.7%, MSK<1.2%, FA<3.8%; global R^2^: MSK>0.93; MD>0.90; FA>0.77), confirming relocation robustness for diffusion-derived microstructural markers. Resting-state functional networks displayed high cross-site reproducibility (ICC>0.90), while task-based fMRI activation peaks retained acceptable spatial and magnitude agreement (ICC=0.30-0.60) despite intrinsic physiological variability. In the absence of software or hardware upgrades, physical scanner relocation leaves qMRI biomarkers remarkably intact—providing the empirical foundation required to preserve data integrity across major facility transitions and multi-site longitudinal cohorts.

**Key points that summarize the paper:**

- Hardware transitions do not compromise qMRI data integrity: Physical relocation of a 3T scanner (including field ramp-down, 30 km transport, and re-shimming) introduces no greater variance in quantitative MRI metrics than baseline within-site test–retest assessments.
- Multimodal biomarker robustness: Brain morphometry, T1 relaxometry, diffusion tensor and kurtosis, resting-state functional connectivity display comparable within- and cross-site stability reflected by different metrics (percent variation, intraclass correlation coefficient and Spearman correlation)
- Empirical backing for longitudinal research: In the absence of hardware or software upgrades, multimodal qMRI biomarkers remain remarkably stable, reassuring researchers that long-term cohort studies and multi-site harmonization can survive major facility relocations.

**Graphical Abstract:** This study evaluates the impact of physically relocating a 3T MRI scanner on neuroimaging biomarkers. By comparing before and after relocation sessions on healthy volunteers, we conclude that the procedure does not introduce significant variability. Parameters for morphometry, T1 relaxometry, diffusion, and resting-state functional connectivity show high stability. Task-based fMRI showed greater within-site variability, but no increase in variability following relocation, consistent with the inherently higher variability of task-evoked responses.. The results confirm the integrity of qMRI data after infrastructure relocation, ensuring reliability for longitudinal studies.

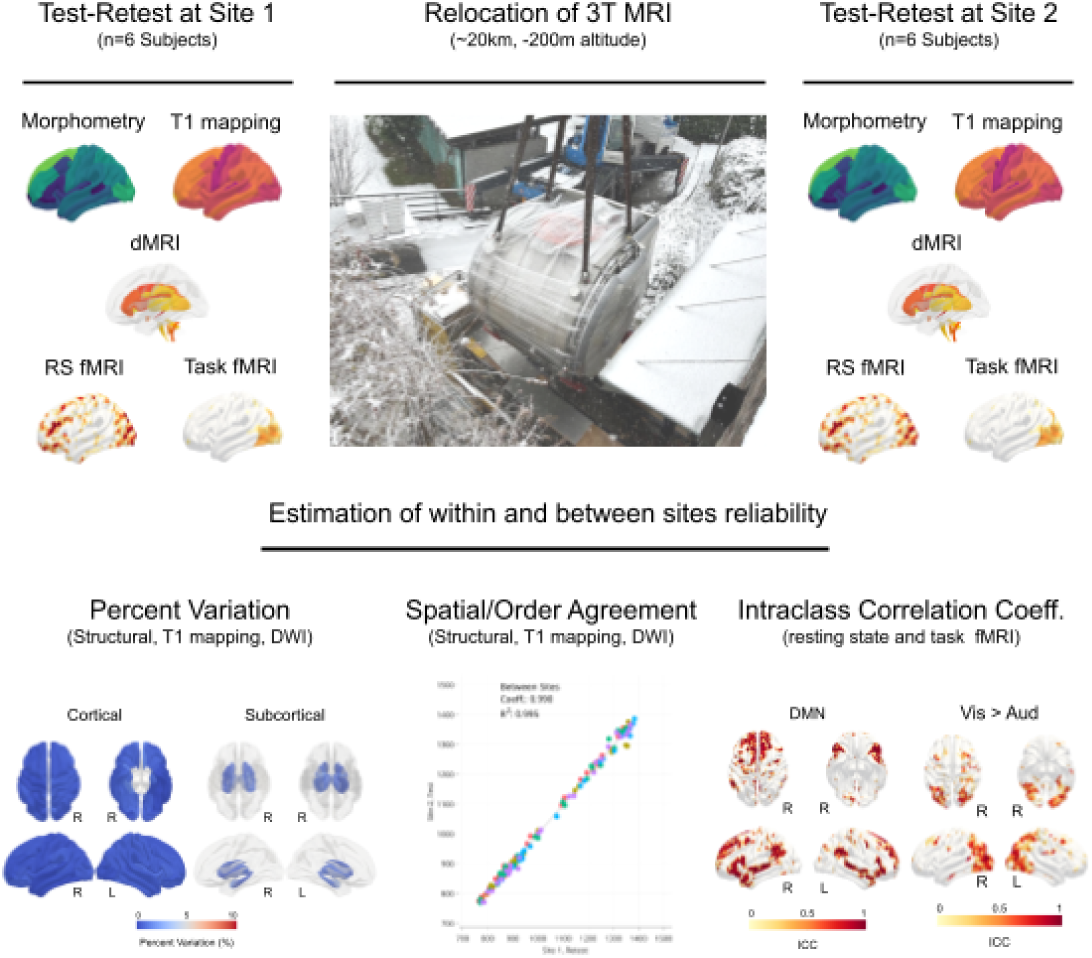

## Introduction

Human brain quantitative magnetic resonance imaging (qMRI) is increasingly relied upon to derive reproducible, biologically meaningful measures of neural structure and function, underpinning longitudinal studies of development, aging, and neurological disease [Váša 2025; Buonincontri 2019]. These metrics—including morphometric, diffusion, relaxometric, task and intrinsic functional measures—are now widely used as candidate biomarkers for subtle brain changes in health and disease [Saltarelli 2025]. Their utility, however, critically depends on the stability of derived measurements across time and experimental conditions. A particularly challenging, though relatively uncommon, scenario is MRI scanner relocation, in which an entire system is physically transferred to a new facility. Even when neither hardware nor software changes are intentionally introduced, relocation can introduce changes in ambient conditions, power and cooling infrastructure, radiofrequency shielding, and local electromagnetic environments, all of which may influence signal properties [Buonincontri 2019; Elliot 2020]. Additionally, hardware components may be affected during the relocation process itself (e.g., system ramp-down, transport, and ramp-up). As research centers expand, consolidate, or modernize their imaging facilities, the ability to ensure continuity of qMRI metrics after scanner relocation has become a key methodological and practical concern for longitudinal cohorts, multi-site consortia, and large-scale harmonization efforts. Yet, the impact of scanner relocation on the reproducibility of qMRI-derived measures remains largely unquantified.

Understanding whether scanner relocation introduces meaningful variability requires placing it in the context of established evidence on qMRI reproducibility across sessions, scanners, and populations. Structural morphometric measures from T1-weighted MRI—such as cortical thickness and regional gray matter volume—generally show high test–retest reliability and long-term stability, supporting their use in longitudinal studies [Reuter 2012; Zuo, Xing 2014] and in applications such as lesion segmentation [Ribaldi 2021]. Diffusion tensor metrics such as fractional anisotropy (FA) and mean diffusivity (MD) also demonstrate good reproducibility when acquisition protocols are harmonized, although variability increases for higher-order diffusion models such as diffusion kurtosis imaging [Kochunov 2015]. Quantitative relaxometry, including T1 and T2 mapping, has likewise shown good repeatability, though systematic biases can arise across vendors and implementations [Bojorquez 2017; Boudreau 2024]. In contrast, functional MRI—particularly resting-state connectivity—exhibits more variable reliability, with reproducibility strongly dependent on preprocessing choices and signal-to-noise characteristics [Zuo, Xing, 2014; Jovicich 2016; Noble 2019]. To our knowledge, only one prior study has directly examined the impact of MRI scanner relocation. Melzer et al. [Melzer 2020] demonstrated that brain morphometry and diffusion tensor metrics acquired before and after relocation of a 3T scanner remained highly consistent, with negligible relocation-related variability when protocols and processing pipelines were kept constant. The growing number of methodological and multi-scanner reliability studies citing this work underscores the field’s need for empirical evidence that qMRI biomarkers remain valid across major hardware transitions.

In the present study, we expand these prior findings by evaluating the effects of scanner relocation across a substantially broader range of quantitative MRI modalities. We examined data acquired before and after the transfer of a whole-body MRI system from a decommissioned area of our research center to a newly constructed facility approximately 30 km away and differing by ∼100 m in altitude. Specifically, our work extends prior relocation studies in three main ways: (1) we assess a different 3T scanner vendor; (2) we directly compare within-site test–retest reliability at both locations with across-site (pre/post-relocation) reliability; (3) beyond morphometry and diffusion tensor imaging (DTI), we evaluate relocation effects on T1 relaxometry, diffusion kurtosis, resting-state network connectivity, and task-based functional networks. Together, this framework provides a comprehensive assessment of the robustness of qMRI-derived biomarkers to scanner relocation and informs the continuity of longitudinal neuroimaging studies across changing experimental environments.

## Materials and Methods

Data were acquired on a 3T MRI scanner (MAGNETOM Prisma, Siemens Healthcare, Forchheim, Germany) equipped with 80mT/m gradient strength and 200T/m/s of maximum slew rate; MR signal was collected with a 64-channel phased-array receive coil for head and neck, software version Syngo MR E11. Six volunteers (mean age= 31 ± 6 years, 3 females) without neurological and/or psychiatric disease history gave written informed consent to participate in this study, which was approved by the Ethical Committee of the University of Trento, Italy. All procedures adhered to the Declaration of Helsinki and applicable data-protection regulations. Each of them performed a test-retest imaging session before and after scanner relocation, for a total of four measurements. The entire set of measurements at the new location was collected within 6 months of receiving the formal authorizations issued by the regulatory bodies. The imaging protocol for each session included: volumetric whole-brain T1-weighted structural imaging, multishell diffusion weighted imaging, resting state BOLD and BOLD functional magnetic resonance imaging (fMRI) during a brain localizer task. Details of each imaging session and strategies for data analysis are reported in the following sections; acquisition parameters are summarized in Table 1.

**Table 1.** Summary of key acquisition parameters for all the sequences employed in the reproducibility study.

| Image Modality | Sequence | Key Acq. Parameters | Acq. Time |
| --- | --- | --- | --- |
| Anatomical | 3D meMPRAGE | TR = 2.53 s, TE = 1.7, 3.55, 5.41, 7.27 ms,<br>TI = 1.1 s, voxel size = 1 mm isotropic,<br>176 sagittal slices, IPAT=2 | 3 min 40 s |
|  | 3D csMP2RAGE | TR = 5.00 ms, TE = 2.88 ms, TI<sub>1</sub>/TI<sub>2</sub> = 0.7/2.5 s, FA<sub>1</sub>/FA<sub>2</sub> = 4°/5°, voxel size = 1.0 mm isotropic, 208 sagittal slices, regularization factors= 6x10<sup>-4</sup>/4x10<sup>-4</sup>, undersampling factor= 4,6x, Denoise Lambda = 5 | 6 min 03 s |
| Diffusion | 4-shell dMRI | TR/TE = 4200/76 ms, voxel size = 2.0 mm isotropic, *b* = 0, 700, 1000, 2850 s/mm<sup>2</sup> (32, 64, 64 dir), Δ/δ = 37.5/14.7 ms, AP/PA phase encoding, IPAT = 6 | 13 min 11 s |
|  |  | For distortion correction: same with reversed phase-encoding, 5 b=0 volumes | 0 min 52 s |
| **Functional**<br>**(Resting-State)** | 2D Multi-Echo<br>Multi-Band BOLD<br>EPI (CMRR) | TR = 1000 ms, FA = 77°, TE = 15.8, 28.1, 40.4 ms, voxel size = 3.8 mm isotropic, GRAPPA factor = 2, SMS factor = 4, 620 volumes, anterior-posterior phase-encoding. | 10 min 40 s |
|  |  | For distortion correction: same with reversed phase-encoding, 5 volumes | 0 min 25 s |
| **Functional**<br>**(Task-Based)** | 2D Single-Echo<br>BOLD EPI | TR = 2000 ms, FA= 65°, TE = 30,4 ms, voxel size = 2 mm isotropic, SMS factor = 3, 170 volumes. | 5 min 40 s |
|  |  | For distortion correction: GRE field maps. TR = 682 ms, TE = 4.92, 7.38 ms, voxel size = 2.0 mm isotropic. | 2 min 19 s |

**Abbreviations:** meMPRAGE = multi-echo MPRAGE, csMP2RAGE = compressed sensing MP2RAGE, TR = Repetition Time; TE = Echo Time; TI = Inversion Time; FA = Flip Angle; BOLD = Blood Oxygenation Level Dependent; EPI = Echo Planar Imaging; GRAPPA = GeneRalized Autocalibrating Partial Parallel Acquisition; SMS = Simultaneous Multi-Slice.

### Structural MRI

For each participant, two high-resolution T1-weighted structural sequences were collected, namely a research application compressed-sensing MP2RAGE (csMP2RAGE, Mussard 2020) sequence and a multi-echo MPRAGE sequence (meMPRAGE, van der Kouve AJW 2008), in order to also enable a direct comparison of morphometric estimates derived from different 3D T1-weighted acquisition strategies.

Both structural sequences were acquired with an isotropic voxel size of 1 mm, providing whole-brain coverage with 208 and 176 slices, respectively. The csMP2RAGE acquisition was characterized by a repetition time (TR) of 5 ms and an echo time (TE) of 2.88 ms, with two different inversion times (TI) of 0.7 and 2.5 s using two different flip angles of 4 and 5 degrees; undersampling factor was set to 4.6x and two regularization values were used for INV1 and INV2, respectively: 6×10^-4^; 4×10^-4^. The meMPRAGE sequence was acquired with a TR of 2.53 s and four TEs of 1.7-3.55-5.41-7.27 ms, matching the spatial resolution of the csMP2RAGE acquisition, with a TI of 1.1 s, GRAPPA acceleration factor: 2. These acquisition parameters were selected to ensure comparable anatomical contrast while preserving the intrinsic signal characteristics of each sequence.

Cortical and subcortical morphometric analyses were performed using the FreeSurfer longitudinal processing pipeline (recon-all, version 7.3.2) [Reuter 2012], which was applied independently to data acquired with the csMP2RAGE and the meMPRAGE sequences. Since each of these sequences offers multiple on-line image reconstruction options, we opted for the most integrated and informative: uniform denoised (UNI-DEN) for the csMP2RAGE; root mean squared echo combination for the meMPRAGE. The longitudinal workflow consists of three main stages. First, each time point for each subject was segmented separately using the standard cross-sectional recon-all FreeSurfer pipeline. Second, an unbiased within-subject template was generated by combining all available time points, and this template was subsequently processed with recon-all to establish a robust anatomical reference. Finally, each individual time point was reprocessed in a longitudinal manner, using the subject-specific template as prior information. The cortical and subcortical volumes of each subject obtained from this final longitudinal stage were then used for the within- and across-site group test-retest morphometry evaluation.

### T1 Relaxometry

The use of csMP2RAGE enabled the fast measurement of whole-brain T1 relaxation time at the same spatial resolution of anatomical scans [Mussard 2020]. Fitting and extrapolation of 3D T1 maps was obtained via the online reconstruction pipeline of the sequence, with no further processing applied afterwards. For each subject, brain parcellations obtained from the morphometric analyses on csMP2RAGE were used to extract the median T1 value from cortical and subcortical ROIs. These averaged T1 values in cortical and subcortical regions were then used for the group test-retest evaluation within- and across sites.

### Diffusion MRI

Diffusion MRI data were acquired using a diffusion-weighted sequence with 2 mm isotropic spatial resolution, n = 75 pure axial slices, echo time/repetition time (TE/TR) of 76/4200 ms and GRAPPA (factor = 2) plus SMS (factor = 3), for a total acceleration factor PAT = 6, full-space sampling. Diffusion sensitisation was implemented across four shells in the anterior-posterior phase encoding direction (b = 0, 700, 1000, and 2850 s/mm²), with 32, 64, and 64 gradient encoding directions for the non-zero *b*-values, respectively, and Δ/δ=37.5/14.7 ms. A separate posterior-anterior phase-encoded direction acquisition with 5 *b* = 0 volumes was acquired for distortion correction.

Pre-processing of diffusion data was carried out using a combination of established software tools [MRtrix3, https://www.mrtrix.org; FSL, https://fsl.fmrib.ox.ac.uk/fsl/docs]. Thermal noise was first reduced using Marchenko–Pastur principal component analysis denoising as implemented in MRtrix (dwidenoise). Gibbs-ringing artefacts were subsequently corrected using mrdegibbs from the same software suite. Susceptibility-induced geometric distortions were addressed using FSL’s topup, followed by eddy current and motion correction performed with eddy_openmp (FSL version 6.0.3). Finally, intensity inhomogeneities were corrected via bias-field correction using dwibiascorrect (MRtrix); details about the pipeline are provided in [Novello 2022].

Diffusion modeling was performed using the DIPY library [Eleftherios 2014]. DTI was used to derive standard scalar metrics, including FA and MD. In addition, mean signal diffusion kurtosis imaging (MSDKI) was employed to estimate mean signal kurtosis (MSK), providing complementary information on non-Gaussian diffusion behavior.

Region-of-interest (ROI) analyses were conducted using both white matter tract and anatomical atlases. Selected white matter tracts were defined according to the ICBM-DTI-81 atlas [Mori, 2008], while cortical, subcortical, and cerebellar gray matter regions of interest were obtained from the FreeSurfer automatic subcortical segmentation (aseg) atlas. Spatial normalization and registrations were performed using ANTs [Tustison 2021]. For tract-based analyses, individual FA maps were linearly registered to the ICBM-DTI-81 diffusion-weighted standard space at 2 mm isotropic resolution, and binarized atlas segmentations were applied to extract median FA values within each tract. To derive regional MD estimates, linear registrations were computed between each participant’s first *b* = 0 volume and the corresponding T1-weighted anatomical image; these transformations were then applied to the atlas segmentations, and median MD values were calculated for each ROI.

For each subject, these median white matter FA (ICBM-DTI-8 tract atlas regions) and gray matter MD (subcortical FreeSurfer parcellations) estimates were then used for the within-and across-site group test-retest morphometry evaluation.

### Resting-state fMRI

Resting-state data were acquired using a multi-echo multi-band sequence [Moeller 2010] with 3.8mm isotropic resolution, and repetition time/echo times (TR/TEs) of 1000/15.8-28.1-40.4ms with GRAPPA factor 2 and Simultaneous Multi-Slice factor 4.

Data was preprocessed with a custom pipeline based on FSL. Briefly, the preprocessing workflow was executed in three primary phases: spatial coregistration, temporal and spatial preprocessing, and multi-echo combination with denoising.

First, structural T1-weighted images were normalized to the MNI152 space, and the functional EPI volumes were coregistered to the native T1-weighted space using Boundary-Based Registration (BBR) via FSL’s FLIRT, which also facilitated the transformation of anatomical segmentations into the native EPI space. Second, standard preprocessing was applied to the sequence, which included slice-timing correction using custom extracted parameters and rigid-body motion correction utilizing FSL’s MCFLIRT. During this phase, motion outliers were quantified by calculating Framewise Displacement (FD) and DVARS, followed by the generation of preliminary temporal signal-to-noise ratio (tSNR) maps. Finally, the preprocessed multi-echo datasets were processed using TE-dependent analysis (TEDANA) framework [DuPre 2021]. This step optimally combined and applied independent component analysis to denoise the data by separating BOLD-like from non-BOLD-like signals, concluding with the extraction of final tSNR maps for both the optimally combined and fully denoised series.

Denoised time series were then used to compute the group-level ICA decomposition, thus obtaining n=20 spatial maps that were finally injected into a dual regression scheme to derive subject-specific versions of group-level resting state networks (FSL dual_regression). Networks obtained from the two experimental sessions were compared for spatial differences with paired-sample, non-parametric, permutation testing (FSL randomise); t-stats maps were converted to p-value maps to evaluate statistically significant differences. To control for type 1 errors resulting from multiple comparisons, threshold-free cluster-enhancement (TFCE) correction was applied to spatial maps and Bonferroni correction was applied at the resting state network (RSN) level. Labelling of RSNs was done by comparing group level ICAs to Yeo 2011 7 Networks [Yeo 2011] parcellation via Dice Similarity index.

### Task-based fMRI

Task-based fMRI data (see Table 1 for acquisition protocol) were acquired using the fast functional localizer paradigm developed by Pinel and colleagues [Pinel 2007], a validated 5-minute fast event-related protocol widely used for mapping sensory, motor, and cognitive networks through alternating visual and auditory stimuli, left- and right-hand button presses, and calculation and sentence-listening/reading tasks, respectively. For the purposes of the present study, we focused on the sensory and the motor networks, which are the most stable both within and across individuals [Pinel 2007]. Further details regarding stimulus design, timing, and sequence optimization are provided in Pinel and colleagues [Pinel 2007]. Stimuli were generated and presented using PsychToolbox routines (PsychToolbox, http://psychtoolbox.org), running in MATLAB (ver. R2019b, The Mathworks, Inc.), using the same computer and monitor (3840 x 2160 pixels, 60Hz) at both sites. The functional images were preprocessed using the Statistical Parametric Mapping (SPM12, https://www.fil.ion.ucl.ac.uk/spm/software/spm12/) in MATLAB, including slice-timing correction, realignment of each scan to the middle of each run, coregistration of functional and anatomical image, segmentation and normalization to the Montreal Neurological Institute (MNI) space. An 8-mm FWHM Gaussian smoothing kernel was applied before GLM analyses.

Task-related activity was estimated at the individual level using a general linear model (GLM), which included 9 task regressors corresponding to the nine conditions of the Pinel localizer ( the presentation of flashing checkerboards (1), visual and auditory instructions to left-and right- hand button presses (2-5), visual and auditory presentation of sentences (6-7), visual and auditory presentation of subtraction problems (8-9)) and six movement regressors obtained from pre-processing. Task regressors were convolved with the canonical hemodynamic response function. Visual activation maps were obtained from the contrast of all visually presented stimuli versus all auditory presented stimuli, whereas auditory activation maps were obtained from the reverse contrast. Maps corresponding to the left hand movements were obtained from the contrast of all left vs. all right button presses, and the reverse for the right hand movements. The resulting subject-specific contrast maps were used for subsequent analyses.

Both whole brain and ROIs analyses were conducted. We defined sensory and motor ROIs on the basis of the results from an independent sample of 33 healthy adults (mean age = 21.8±2.8 years; 23 F), tested in the context of a different project (University of Trento ethical approval n. 2022-058) with the very same Pinel localizer protocol used here. The sensory ROIs were defined from the group-level l (FWE-corrected clusters) activation maps for the Visual>Auditory and Auditory>Visual contrasts (one ROI per hemisphere). The results substantially replicate those in the original Pinel study [Pinel 2007]: the Auditory > Visual ROI included the superior temporal cortex, including Heschl’s gyrus, the superior temporal gyrus, and adjacent middle temporal regions, while the right hemisphere additionally included the superior temporal pole. The Visual > Auditory ROI included the inferior, middle, and superior occipital gyri, fusiform gyrus, inferior temporal cortex, and superior parietal regions on the left, while the right Visual > Auditory ROI included the inferior and middle occipital and fusiform gyri. The motor ROIs were defined as the press Right > press Left and press Left > press Right contrast, each identifying a contralateral sensorimotor region encompassing the precentral and postcentral gyri (see SI, Figure F1). For each functional ROI, three complementary metrics were extracted from the first-level contrast maps for each participant and session: (I) activation intensity, (II) activation peak location and (III) activation extent. Five participants were included in the analyses; one participant was excluded because he missed one session. Each participant was scanned at two sites (Site 1 and Site 2), with one test and one retest session at each site.

I. Activation intensity was computed as the average contrast estimate across all voxels.
II. Peak location was defined as the 3D spatial coordinates of the voxel exhibiting the highest t-value. In order to assess the spatial stability of peak location across sessions, we computed the Euclidean distance between homologous peak coordinates in the left and right hemispheres were then computed as:

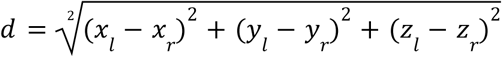

where x_l_,y_l_,z_l_, and x_r_,y_r_,z_r_ correspond to the coordinates of the left- and right-hemispheric peak voxels, respectively. The advantage of this measure is that it removes the potential realignments / normalization variations across sessions and removes the variable “hemisphere” from subsequent statistical analyses, thus halving the number of tests.
III. Activation extent was defined as the number of voxels within each ROI exceeding a statistical threshold of T>3.1 (corresponding to p<0.01).

### Test-Retest Metrics

To define the reliability of the above mentioned measurements, we compared three test-retest sessions:

1) within-site 1, hereafter wSite1 (before relocation), contrasting site1 test vs. site1 retest;
2) within-site 2, hereafter wSite2 (after relocation), contrasting site2 test vs. site2 retest;
3) between-sites, hereafter bSites, defined as site1 retest vs. site2 test, the two closest-in-time across sites sessions.

The test-retest reliability was assessed using four estimators: (I) Percent Variation (PV), representing the increase or decrease between two values relative to their average [Jovicich 2013]; (II) Two-way ANOVA with Session X Site interaction; (III) correlation analysis across sites and sessions; (IV) ICC(3,1) to assess consistency between sessions [Elliot 2020]. Metrics derived from structural scans (3D T1-weighted, T1 maps and diffusion-weighted data) were processed with a ROI-based approach using FreeSurfer cortical parcellations and subcortical segmentations. Intrinsic connectivity and task-based fMRI data were both tested with a combined functional ROI and whole brain approach. Predefined ROIs (group-level RSNs or validation ROIs, respectively) were used to extract voxel-wise metrics (strength of connectivity in the former, strength and extent of response in the latter) within and across sites. Then: ICC(3,1) voxel-wise maps were obtained from z-stat values for each of the considered RSNs; concerning task-based fMRI data, voxel-wise ICC(3,1) maps were generated from first-level contrast estimates to characterize the reliability in terms of spatial distribution of differential activation (contrasts) across the whole-brain. Also in this case, reliability maps were computed separately for each contrast. Additionally, average within-network connectivity, activation strength, number of activated voxels and euclidean distance were processed to obtain network- or contrast-specific estimates of PV. Correlation analysis was consistently performed on each image-specific metrics to compare sessions and sites.

In the following sections we report and discuss results and visuals about the between-sites comparison for all the selected metrics; within site data and correlation analyses are available to the interested reader in the Supplementary Materials. A compact summary of the experimental design, including planning, sequences evaluated and results is reported in Table T1 in Supplementary Materials.

## Results

Figure 1 shows, for a sample subject, a representation of the various metrics (morphometry, relaxometry, diffusion and functional) derived from each imaging modality, for which scanner relocation reproducibility was evaluated. The main finding is that site relocation did not significantly affect any of the metrics evaluated. In what follows, we report the quantitative results for each imaging modality.

**Figure 1.**
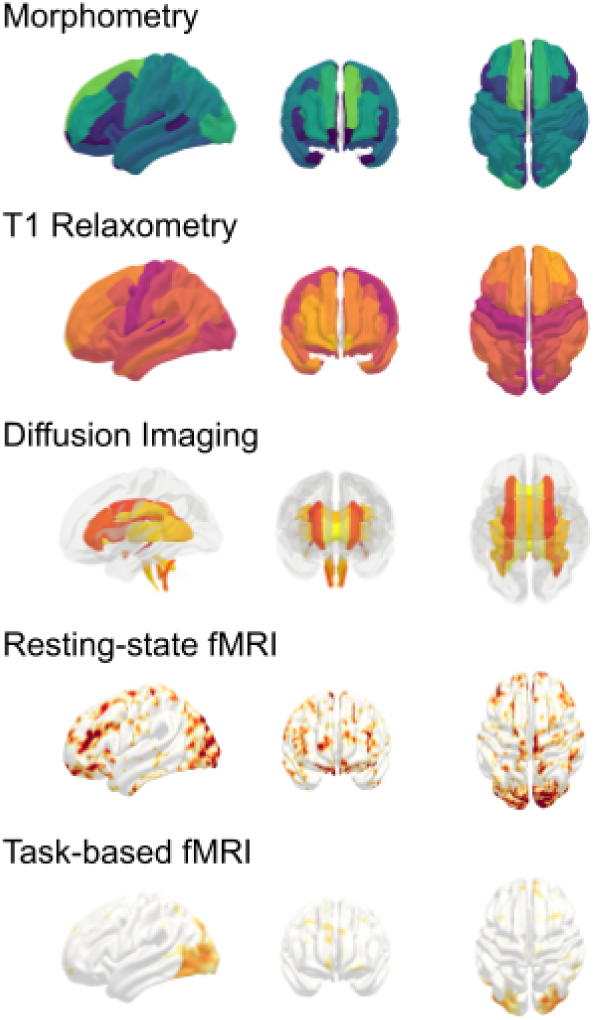
Representative surface-based projections of the different metrics analyzed across sessions and sites at the single subject level.

### Site relocation reproducibility of brain morphometry

*(Figure 2):* Brain morphometry segmentations derived from both csMPR2AGE and meMPRAGE anatomical sequences demonstrated high and similar within- and across-site reproducibility in all evaluated cortical and subcortical regions. In terms of variance and interactions, two-way ANOVAs revealed no significant main effects for Session or Site, nor any interaction effects for either sequence. Focusing on the main effect of PV across sites, group PV remained consistently below 5% across the brain, with whole-brain averages below 1% (csMP2RAGE: 0.86±0.44%; meMPRAGE: 0.84±0.26%; Figure 2). In terms of reliability, ICC revealed excellent reliability for both sequences (avg ICC=0.99). Correlation analysis reported excellent agreement with global R^2^>0.9. Within-site test-retest comparisons yielded similar results (Supplementary Figures A1–A3).

**Figure 2.**
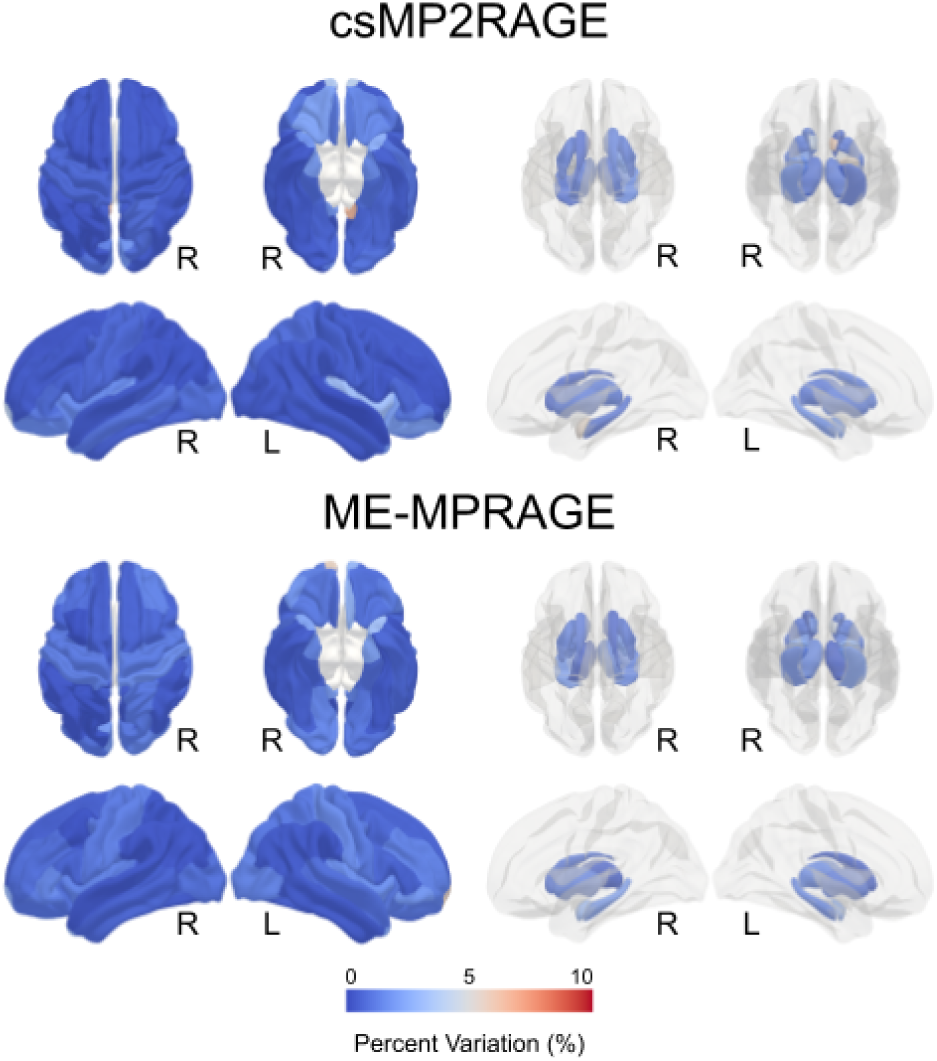
Group averaged PV across relocation sites for cortical (left) and subcortical (right) volumes. At global level, whole brain evaluation shows changes below 5%.

### Site relocation reproducibility of brain T1 relaxometry

*(Figure 3):* Two way ANOVA computed from high-resolution csMP2RAGE T1 maps showed no significant differences both for main effects (Session, Site) and their interaction, in any of the considered ROI. PV across sites remained below the 5% threshold between-sites (Figure 3), with a whole-brain average PV below 1% (0.66±1.6%. ICC(3,1) revealed excellent reliability (average ICC=0.90, R^2^>0.9).

**Figure 3.**
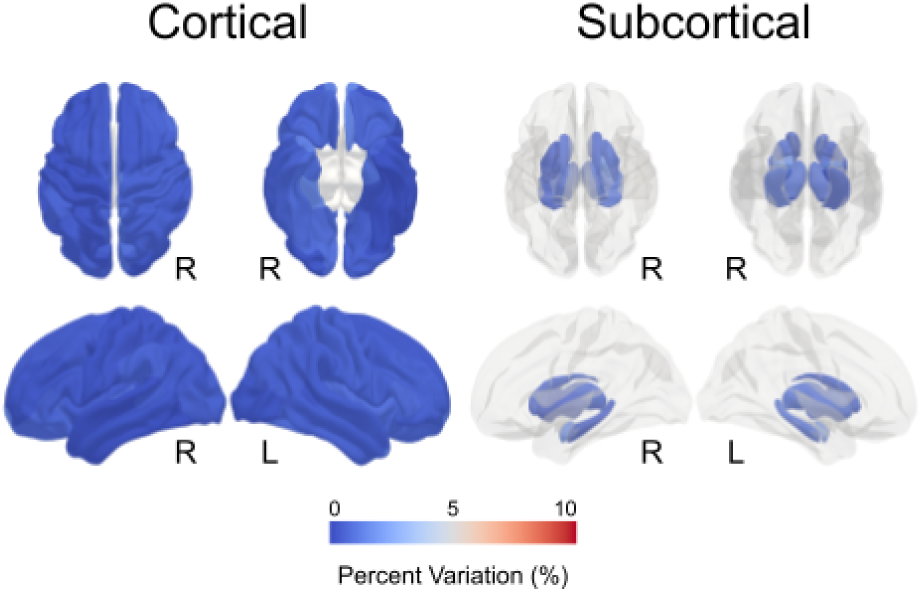
Group averaged PV across relocation sites for quantitative T1 mapping derived from csMP2RAGE for cortical (left) and subcortical (right) brain structures. The between site variation is robustly set below the 5% for the entire brain.

### Site relocation reproducibility of brain microstructure

*(Figure 4).* Two way ANOVA reported non-significant results for both main effects nor their interaction. Relative PV across relocation sites in MD and MSK metrics was less than 5% and demonstrated excellent reliability (MD: PV=1.66%, ICC(3,1)=0.91; MSK: PV=1.18%, ICC(3,1)=0.97). A moderate increased variation of FA was found in ventral and posterior bundles, with values ranging from 5 to 10%. Nonetheless, excellent reliability is preserved across sites (FA ICC(3,1)=0.97), and across the whole brain (global FA PV=3.8%, R^2^>0.9).

**Figure 4.**
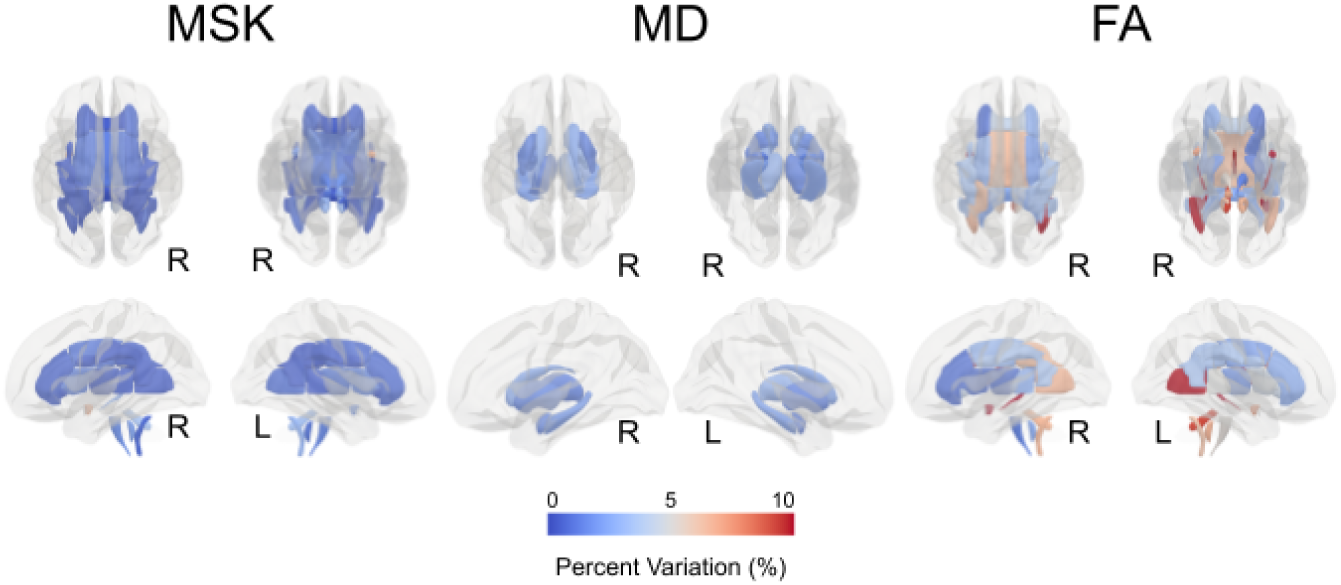
Group PV in diffusion MRI metrics across relocation sites. Microstructural (MSK) and tensor-based values of mean diffusivity (MD) and fractional anisotropy (FA) are evaluated in white matter tracts and subcortical gray matter nuclei. Most of the considered ROIs exhibit a <5% variation, with FA the only exception (<10%) in posterior and ventromedial fiber bundles.

### Site relocation reproducibility of intrinsic functional connectivity

*(Figure 5).* Resting-state connectivity analysis on average Z-score revealed no significant differences for both main effects and their interaction in a two-way ANOVA across all the considered networks. PV of voxel-wise Z-score in each of the RSNs here evaluated ICC(3,1) showed excellent agreement with an estimate of 0.99. Spatial distribution of networks was also tested: non-significant differences were reported with TFCE-corrected non-parametric permutation testing.

**Figure 5.**
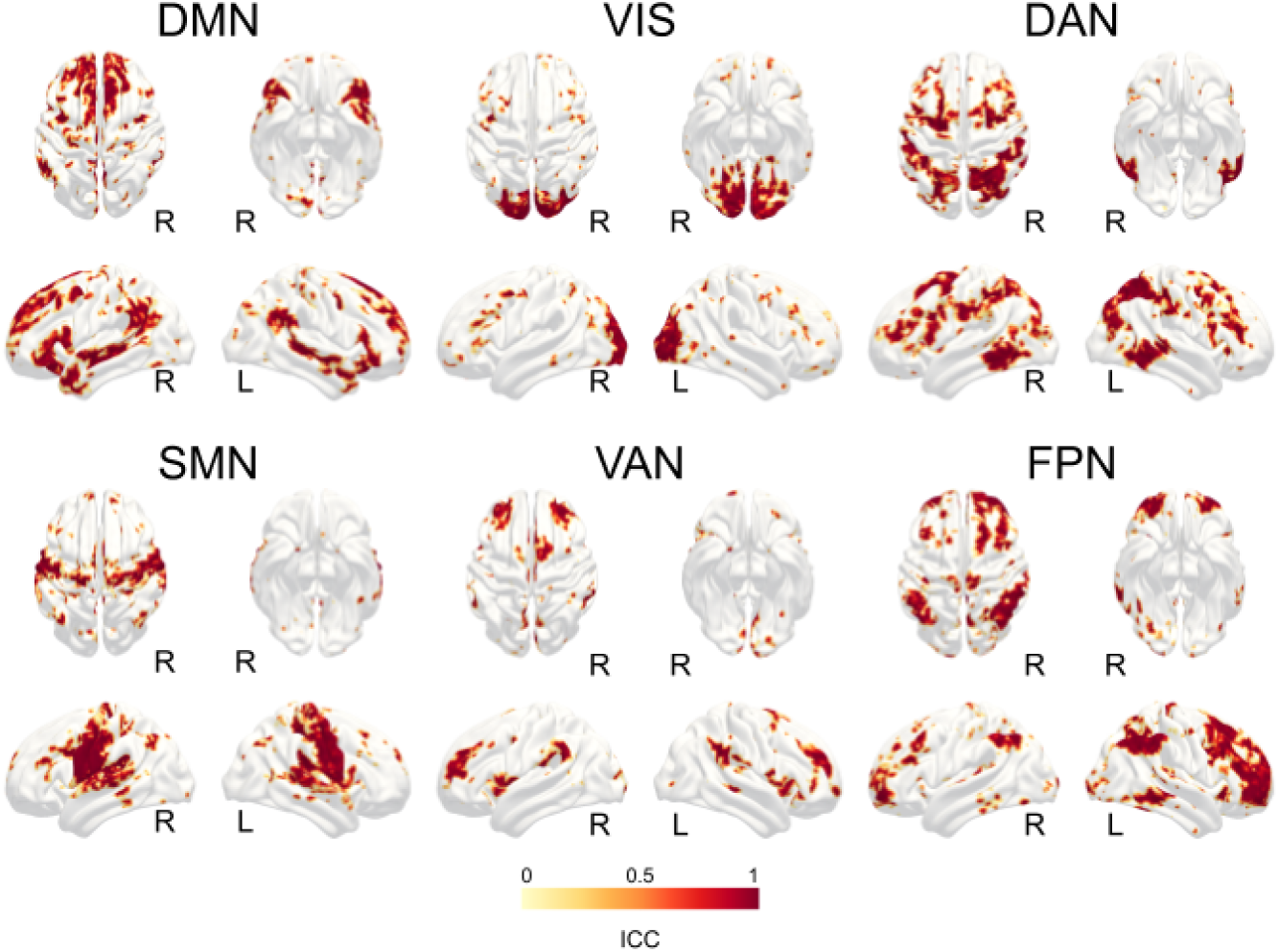
Reproducibility of intrinsic connectivity across scanner relocation sites. Within-network strength in all the major resting state networks is robust when compared before and after relocation. Voxelwise ICC mapping shows an excellent group-level topological agreement between repeated scans. Abbreviations: DMN, default mode network; VIS, visual network; DAN, dorsal attention network; SMN, somatosensory motor network; VAN, ventral attention network; FPN, fronto-parietal network.

### Site relocation reproducibility of task-based Fmri

*(Figure 6).* The strength of neural activation, number of activated voxels and Euclidean distance in the left and right hemispheres, computed separately for the visual and auditory contrasts, revealed no significant main effects (Site, Sessions) nor interaction when tested with a two way ANOVA. In all the considered metrics, the PV strongly exceeded the 5% threshold in all the above mentioned metrics and sessions, a condition confirmed by global ICC values around or below zero. The ICC(3,1) analysis of audio-video and left-right contrasts was more informative. When comparing magnitudes of contrast, a fair reliability for the auditory-visual contrast (ICC(3,1) Aud>Vis = 0.44; Vis>Aud = 0.43), and poor-to-fair reliability for the left-right contrast (ICC(3,1) Right>Left = 0.38; Left>Right = 0.39) is consistently retrieved between sites.

**Figure 6.**
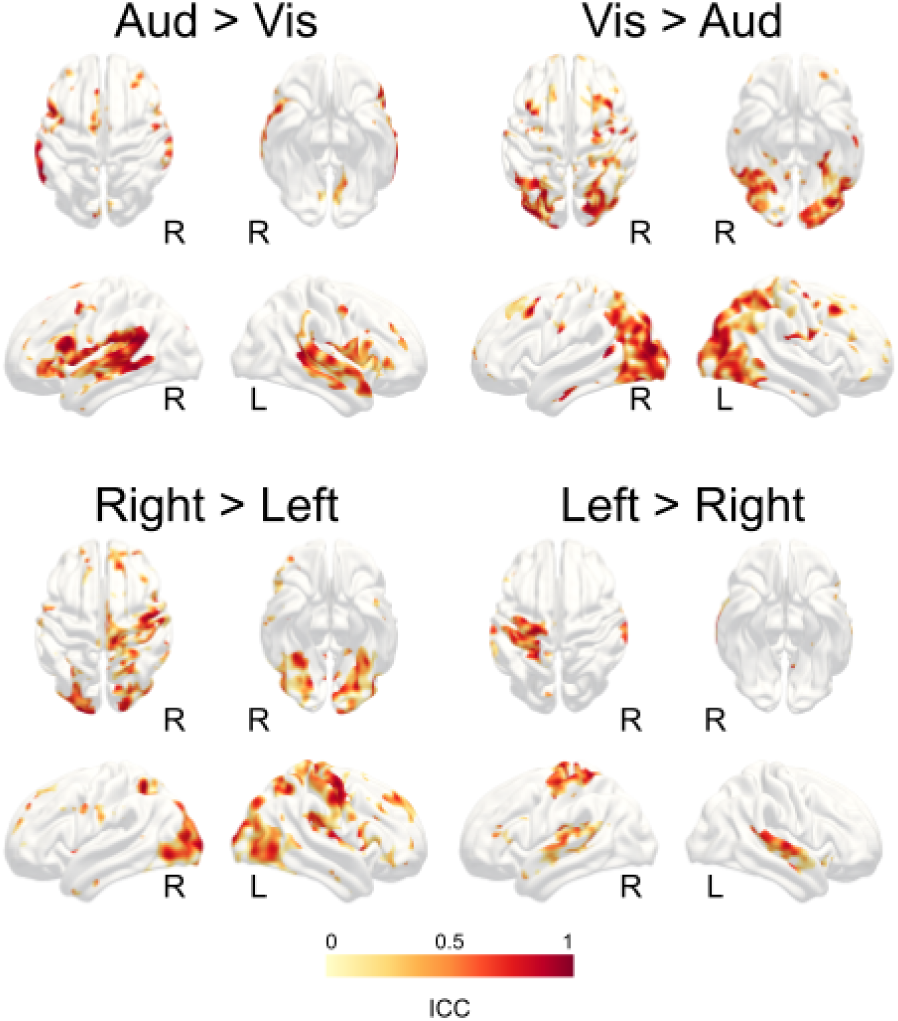
Spatially resolved reliability of activation strength for audio-visual and left-right contrasts, computed across sites. ICC values are in the fair range and poor-to-fair range, respectively.

## Discussion

Relocating a high-end MRI scanner involves a series of invasive technical procedures—from ramping down the static magnetic field, disconnecting power, cooling, and signal lines, and physically transporting the unit, to rebuilding the Faraday cage, ramping up the main field, and re-shimming. To evaluate whether such major infrastructural disruptions introduce systematic biases into derived neuroimaging markers, we conducted a systematic pre- and post-relocation assessment in a cohort of healthy volunteers. Overall, test–retest multispectral measurements demonstrated that relocation had a negligible impact on data consistency across all evaluated metrics. Crucially, reproducibility metrics were equivalent across all three conditions—within original site, between sites, and within new site—confirming that scanner transfer introduces no greater variance than standard within-site test–retest assessments.

Measurements of brain morphometry exhibited test-retest variations below 5%, together with good to excellent reliability (global PV<1%, global ICC>0.99, global R^2^>0.9). Other works in the field [Quattrini 2020; Melzer 2020] reported similar ICC values, although under different testing conditions and pulse sequences, suggesting that scanner relocation does not introduce significant biases in volumetric brain scans. It was far from established that this holds true for two very different pulse sequences in terms of k-space sampling and duration, confirming that different and non-commercial methods (meMPRAGE and csMP2RAGE) are robust, with high SNR and overall excellent quality. In addition, csMP2RAGE provides fast and robust quantitative T1 mapping by fitting the images derived from the two preparation modules. The above-mentioned maps were tested for reproducibility in our sample via ROI based segmentation. Even in this case, PV was very limited, with excellent reproducibility (PV<1%, ICC=0.89, R^2^>0.9). The morphometric and relaxometric dimensions investigated here, quantitative by definition, mark an important reference point for basic neuroscience and clinical investigations addressing brain atrophy and tissue degeneration [Saltarelli 2025].

Similarly, diffusion-derived measures showed low variability, with most of white matter and deep gray matter regions changing less than 5% in tensor-based and kurtosis-derived microstructural assessment across sites and sessions (global PV<2.5%, global ICC>0.9, global R^2^>0.9). Under consistent protocols for acquisition and processing, similar values were reported previously for percent variability [Jovicich 2014, Palacios 2017, Zhou 2018] and ICC [Boekel 2017] across sites and vendors, suggesting that factors other than scanner relocation are responsible for the globally observable variance in white matter. From a clinical perspective, MD and DKI have been successfully exploited in a number of neurodegenerative diseases [Bozzetti 2026], thus ensuring that the longitudinal validity of these metrics after major environmental changes is highly preserved. Implication of these findings lives also in the domain of longitudinal and retrospective assessment of structural connectivity. Extending recent work highlighting diffusion tensor-based structural connectivity as a highly reproducible network proxy [Lizarraga 2025], our results provide further evidence that this metric is also remarkably robust to physical scanner relocation.

Complementing these microstructural findings, we observed a similarly high level of reliability when evaluating intrinsic functional connectivity, both within- and across-sites. Reproducibility of ICA-derived RSNs provided excellent results, with PV<2.5%, ICC>0.9, even better than previously reported [Guo 2012]. In particular, with an interpretation grounded on the same work, these analyses can serve as short- and long-term evaluations on reproducibility of intrinsic functional connectivity. Within-site data are proxies for short-term reproducibility (one week between sessions); between-site comparisons, separated by several weeks - the timeframe needed to fulfill technical and regulatory requirements to resume full operation at the new site - might provide information on longer-term reproducibility. Previous work [Wang JH 2011] reported similar ICC values for both short- and long-term cases; our findings confirm and extend those observations with higher mean ICC values in both within- and between-sites conditions across the major resting state networks. From early attempts in cortical mapping [Orringer 2012] to accurate presurgical planning [Moretto 2024] and prognostic evaluation [Saviola 2022], intrinsic functional connectivity has gained wide popularity in longitudinal assessment of connectivity changes, and its robustness against several potential confounding factors testify in favor not only of reproducible research, but also of advanced clinically relevant applications.

The last dimension considered here, task-based fMRI, opens the discussion towards non-trivial aspects of reproducibility. Overall, from a purely analytical perspective, our data suggests low consistency between sessions, congruent with other studies. There is an ongoing debate on the reliability of results from fMRI that is still far from a consensus [Bennet 2010, Elliot 2020] after more than 10 years of experiments and evaluations, and in this context, our data are aligned. It is a well-established piece of evidence in the field that many confound factors concur in the apparently poor reproducibility of fMRI. Some of them are psycho-physiological: arousal and attentional state, competing cognitive processes, circadian rhythms [Hodkinson 2014]; some derives from the subject’s compliance: caffeine or alcohol intake, drowsiness, and more [Clement 2018]; but all of them inevitably influence the short-term homeostasis of brain perfusion. Some factors are purely technical and very common (signal-to-noise ratio, thermal noise, environmental conditions); others are very rare, and so we consider them negligible for the purpose of this study. Hence the choice of exploiting a multimodal paradigm [Pinel 2007] with documented within-subject robustness between sessions. In agreement with the observation of Pinel, we found that purely geometric features of activations (anatomical locations, extent of the activation and relative distance) are strongly influenced by subject specific factors. What is more interesting is the evaluation of the global quality of activation, e.g. the differential magnitude of functional response to selective paradigms. By comparing the between site data of our study, we found satisfactory agreement in all the considered contrasts (global ICC>0.3), comparable with other works in the field [Elliot ML 2020, Peelen MV 2005]. Although still too far from establishing fMRI as robust and reliable, we can conclude that fMRI data collected under these conditions are minimally affected by a deep restructuring of the experimental environment, like those introduced by relocation of an MR scanner.

At least two major limitations of this study warrant consideration. First, environmental factors—such as precise room temperature, humidity, and electromagnetic shielding integrity—play a critical role in measurement stability across sites. Although baseline environmental controls were maintained by third-party vendors according to manufacturer specifications, historical continuous monitoring logs from the original site were unavailable for direct quantitative comparison due to a data storage failure during the move. To prevent such data loss and account for potential environmental confounds, we have since transitioned to an automated, cloud-based monitoring infrastructure. Based on this experience, we strongly recommend that future relocation or long-term multi-site studies maintain continuous cloud-backed records of ambient conditions alongside routine manufacturer service reports—specifically standard phantom image quality metrics and Faraday cage attenuation logs at the scanner’s operating frequency. Second, we acknowledge that a sample size as low as six subjects might be underpowered to capture more subtle instabilities. This is counterbalanced by the purely paired design and the availability of a full test-retest session at each site. By comparing the three evaluations, we found that between sites reproducibility is no worse than within site’s, a reassuring confirmation about the reliability of the results. The last in line is the choice of the metrics to consider. It would have been interesting to extend the validation towards additional well-established neuroimaging methods with quantitative outcomes, like myelin and susceptibility mapping, brain perfusion with arterial spin labeling or quantification of brain metabolites via single voxel spectroscopy, to name a few. We deliberately prioritized the techniques presented here due to scan time limits and efficiency constraints, under the consideration that the latter are not extensively adopted by the vast majority of researchers who have access to the facility.

To conclude, despite unavoidable technical adjustments and environmental changes associated with a 3T clinical scanner relocation, we observed no significant variation across any of the evaluated neuroimaging metrics. Intraclass correlation coefficients demonstrated high reliability for structural measures (ICC=0.98), diffusion metrics (ICC=0.99), and resting-state connectivity (ICC=0.99), alongside acceptable reliability for task-based fMRI peak localisation (ICC range 0.30–0.60). Together, these findings demonstrate that MRI-derived neuroimaging markers, across a broad range of imaging modalities, remain remarkably robust following scanner relocation (without hardware or software upgrades), reassuring researchers that longitudinal study integrity can be maintained even amidst major infrastructural changes.

## Data Availability

All data produced in the present study are available upon reasonable request to the authors

## Conflict of interest disclosure

The authors have no conflict of interest to declare. Tom Hilbert, Gian Franco Piredda and Tobias Kober are employees of Siemens Healthineers AG.

## Ethics approval statement

The study was approved by the Ethics Committee of the University of Trento, all subjects provided written informed consent.

## Data availability statement

The data and source code used for the analysis presented in this study will be made available upon request.

MRtrix3 - https://www.mrtrix.org/

FSL - https://fsl.fmrib.ox.ac.uk/fsl/docs/

SPM12 - https://www.fil.ion.ucl.ac.uk/spm/software/spm12/

PsychToolbox - http://psychtoolbox.org/

## Supplementary Materials

### Structural MRI

**Fig. A1.**
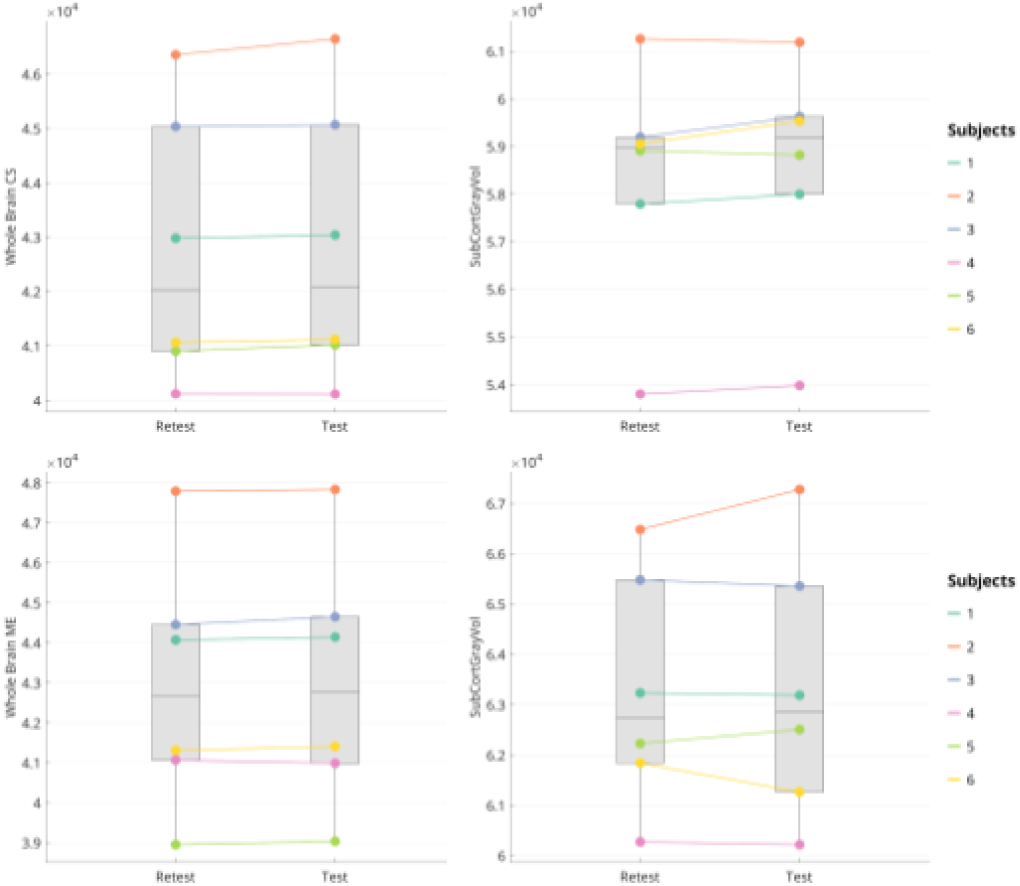
Between sites comparison of subject-level global cortical and subcortical brain volumes

**Fig. A2.**
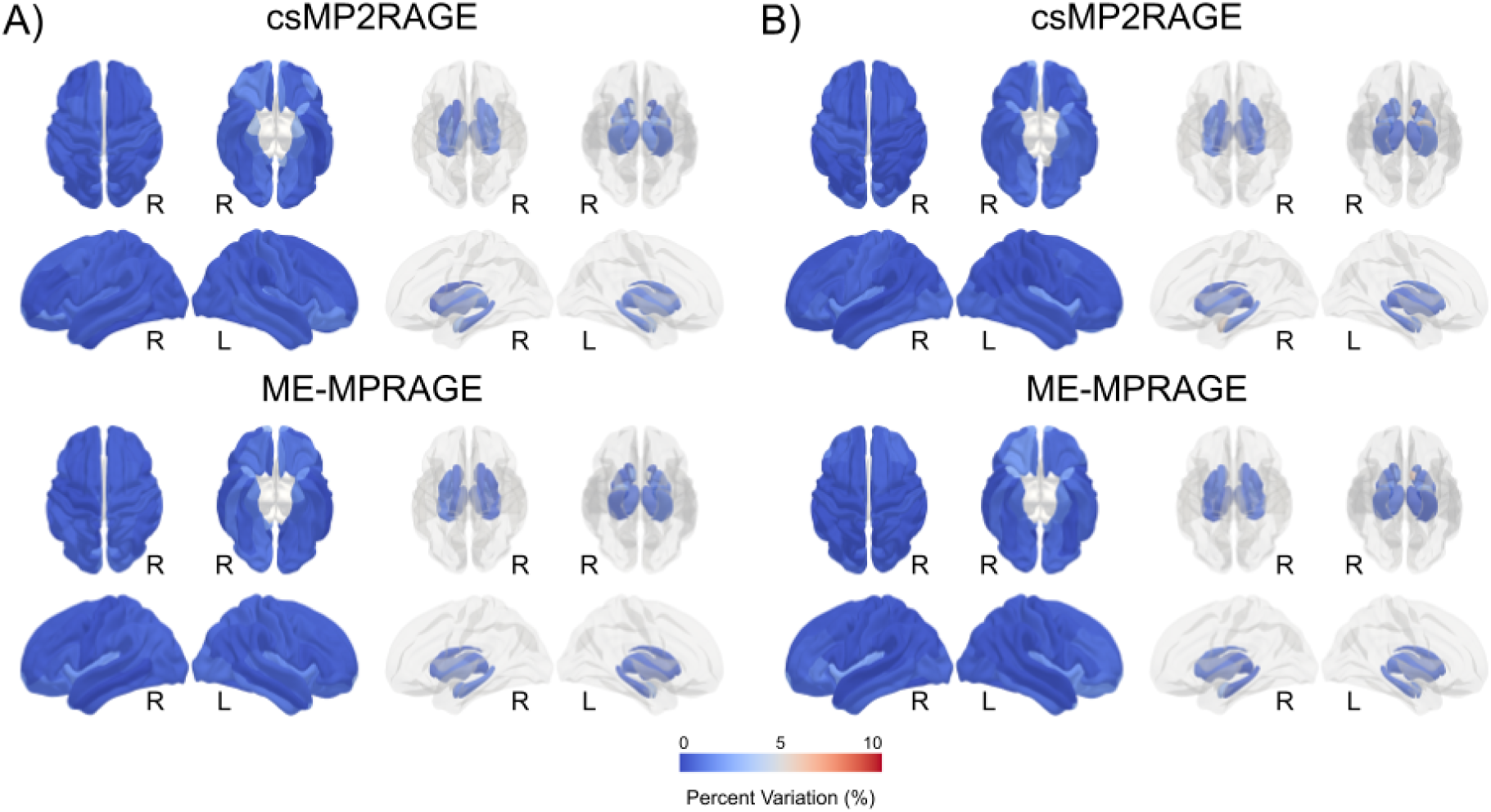
Within site percent variation of cortical and subcortical brain volumes for csMP2RAGE and ME-MPRAGE. Panel A): within Site 1; panel B): within Site 2

**Fig. A3.**
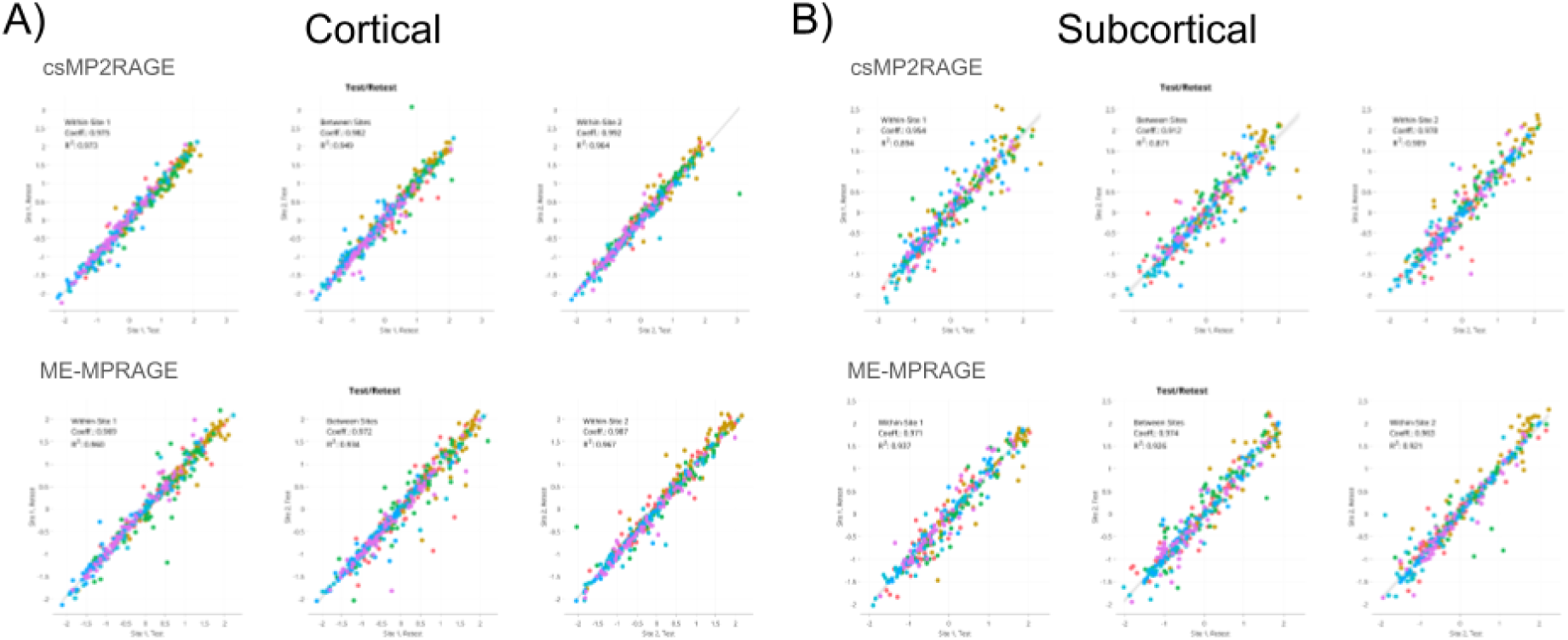
Correlation analysis of test-retest cortical and subcortical volume of individual parcels. Color encodes subjects.

### T1 Relaxometry

**Fig. R1.**
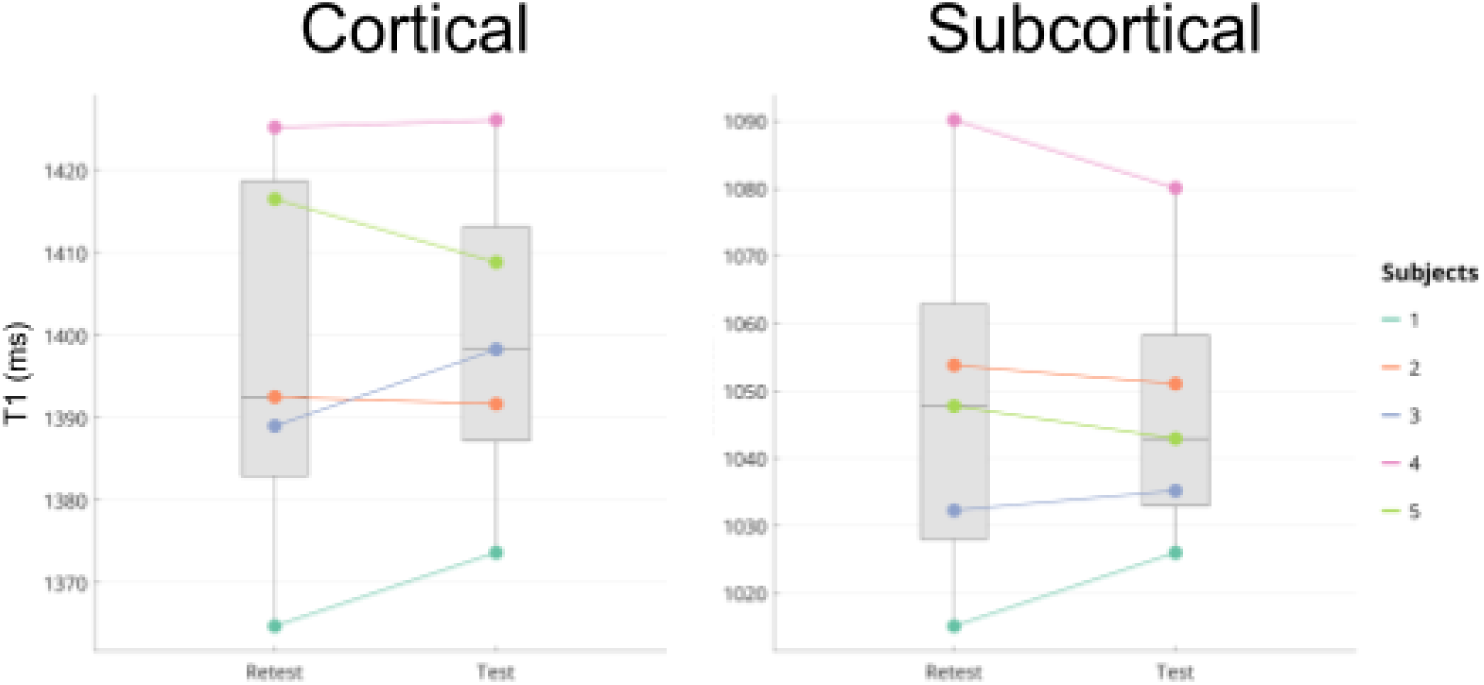
Between sites comparison of subject-level global cortical and subcortical brain T1 relaxation time

**Fig. R2.**
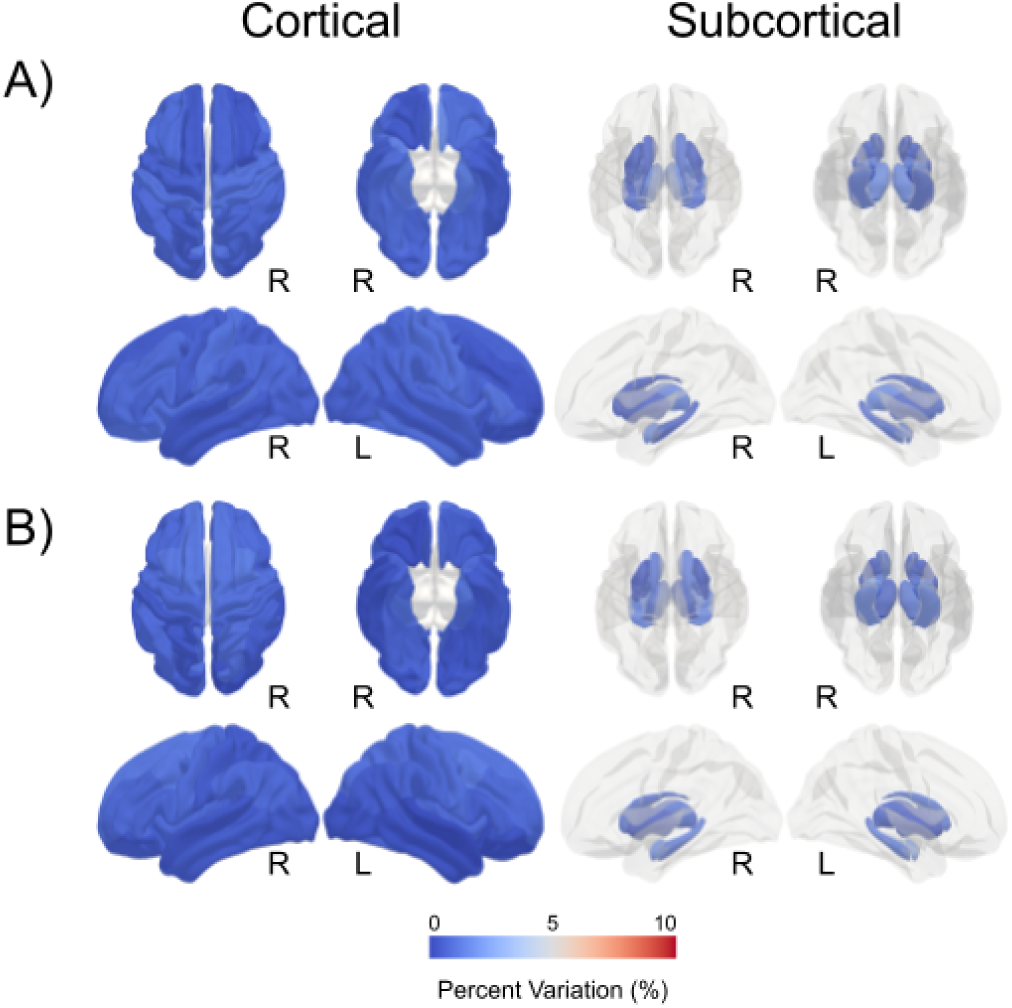
Within site percent variation of cortical and subcortical brain T1 relaxation times fitted from csMP2RAGE data. Panel A): within Site 1; panel B): within Site 2

**Fig. R3.**
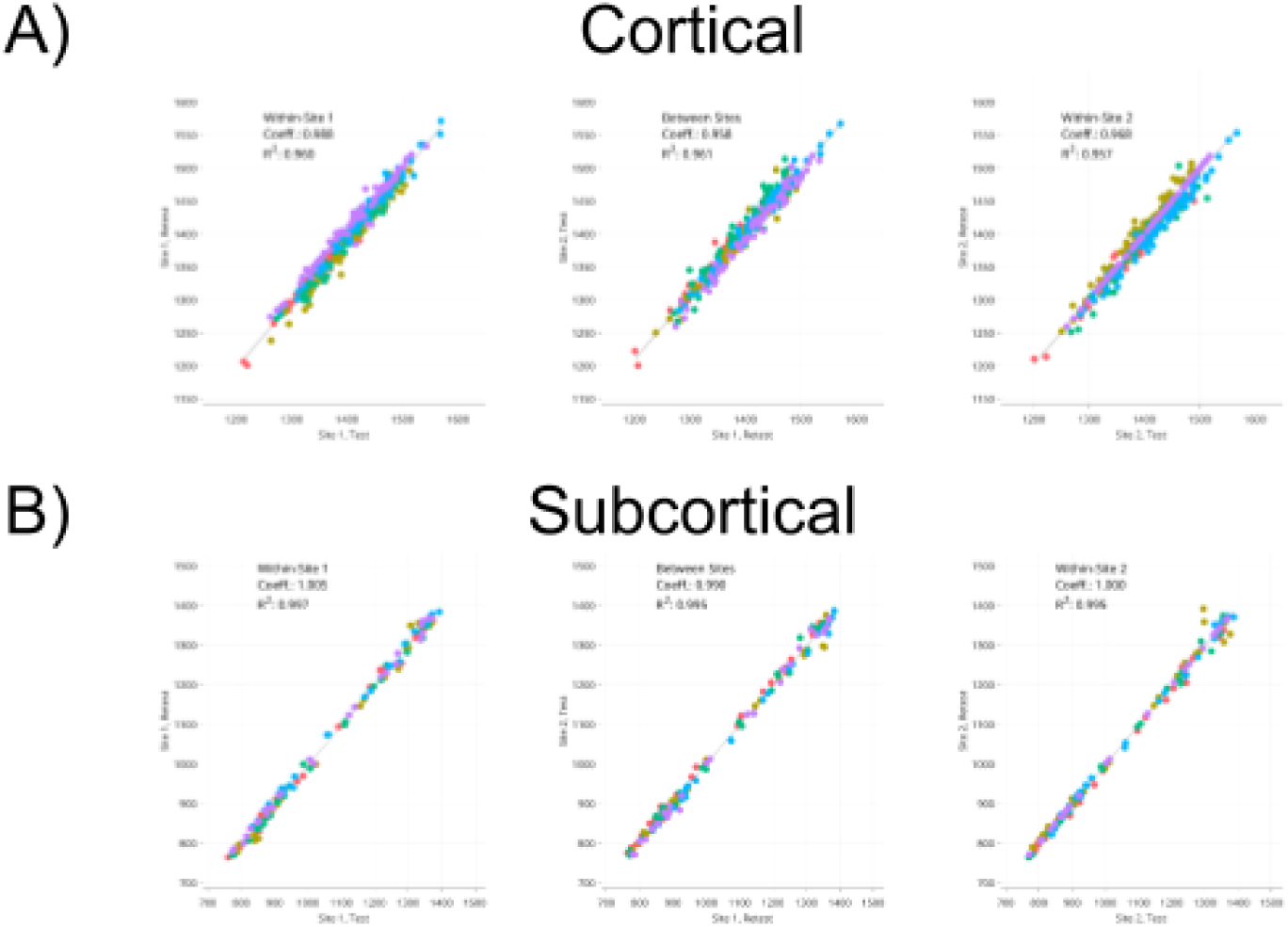
Correlation analysis of test-retest cortical and subcortical T1 relaxation time of individual parcels. Color encodes subjects.

### Diffusion MRI

**Fig. D1.**
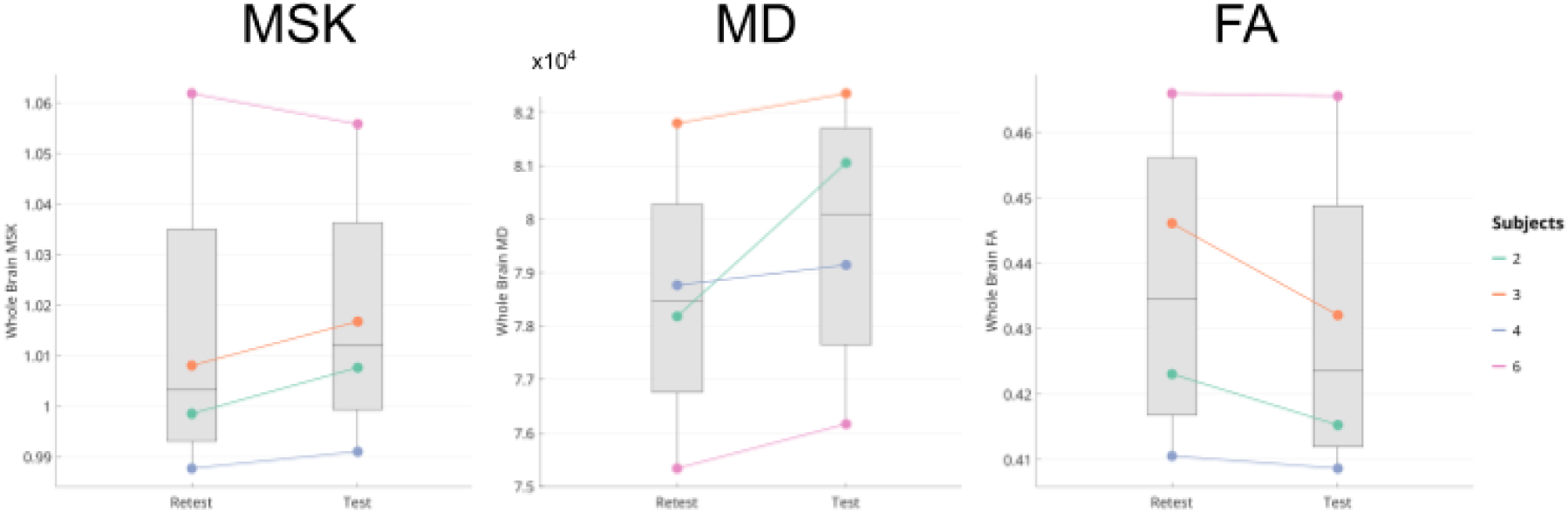
Between sites comparison of subject-level global microstructural and tensor-derived diffusion weighted metrics

**Fig. D2.**
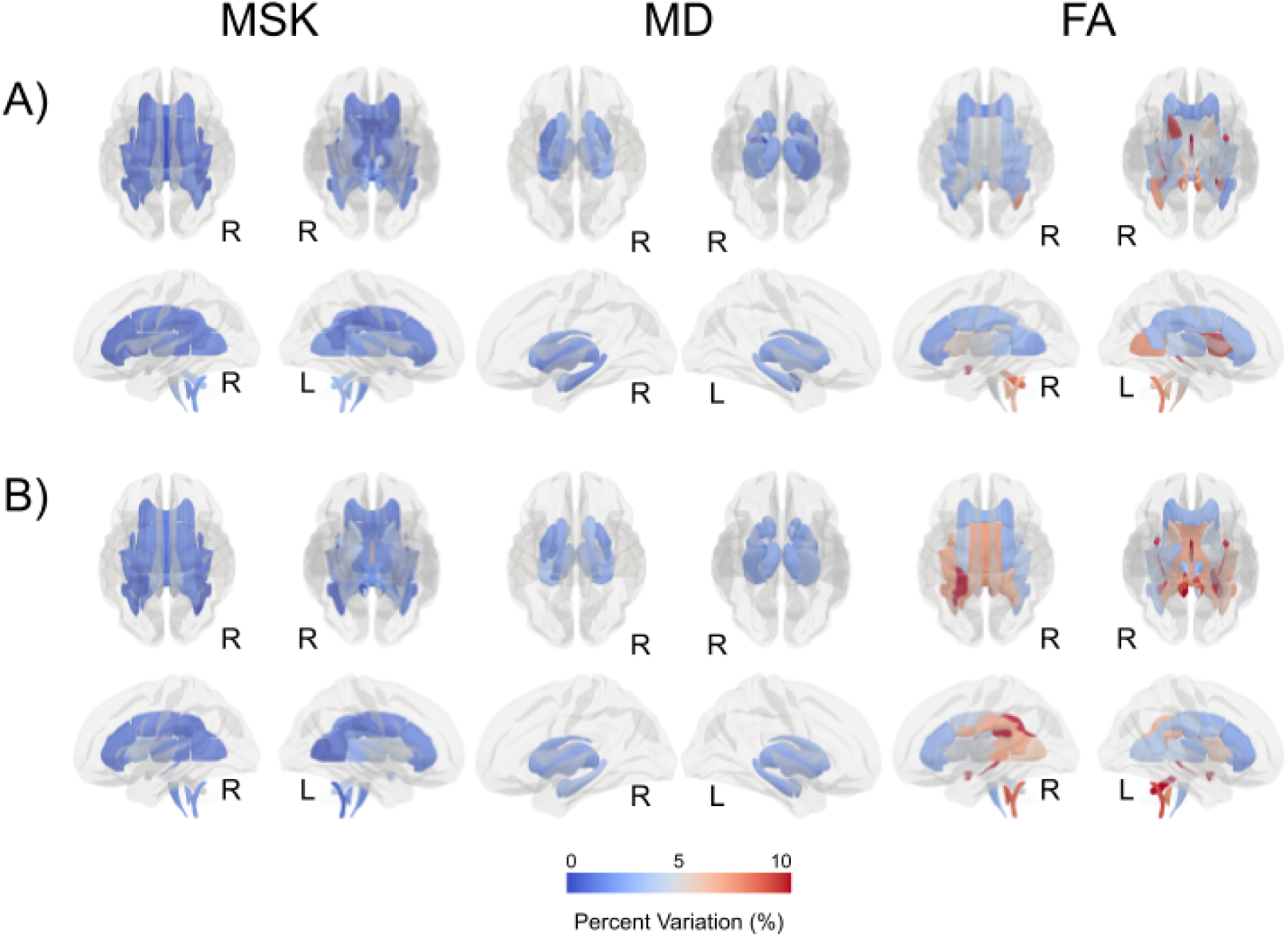
Within site percent variation of microstructural and tensor-derived diffusion weighted metrics. Panel A): within Site 1; panel B): within Site 2

**Fig. D3.**
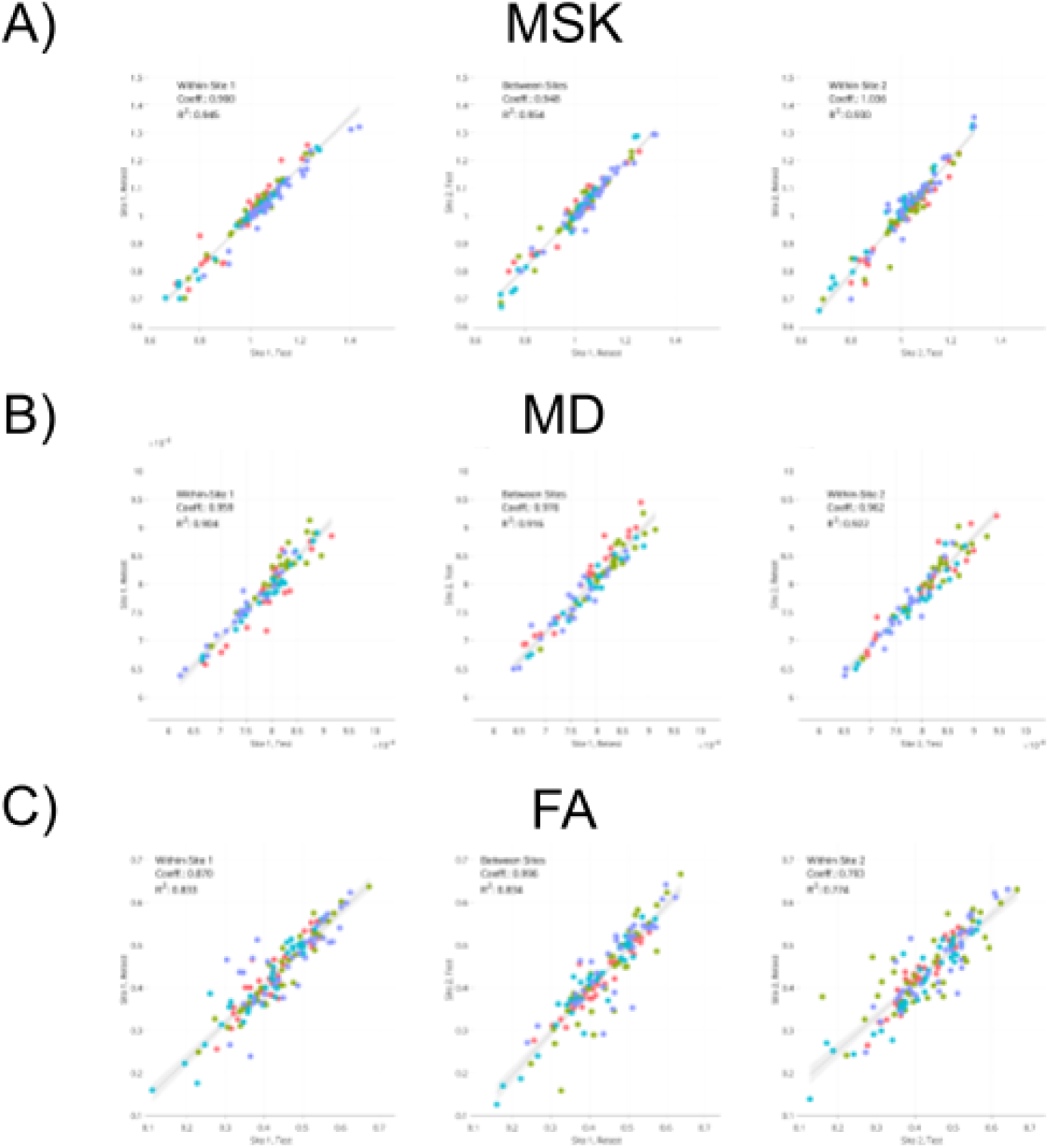
Correlation analysis of test-retest microstructural and tensor-based diffusion weighted metrics of individual parcels. Color encodes subjects.

### Resting-State fMRI

**Fig. C1.**
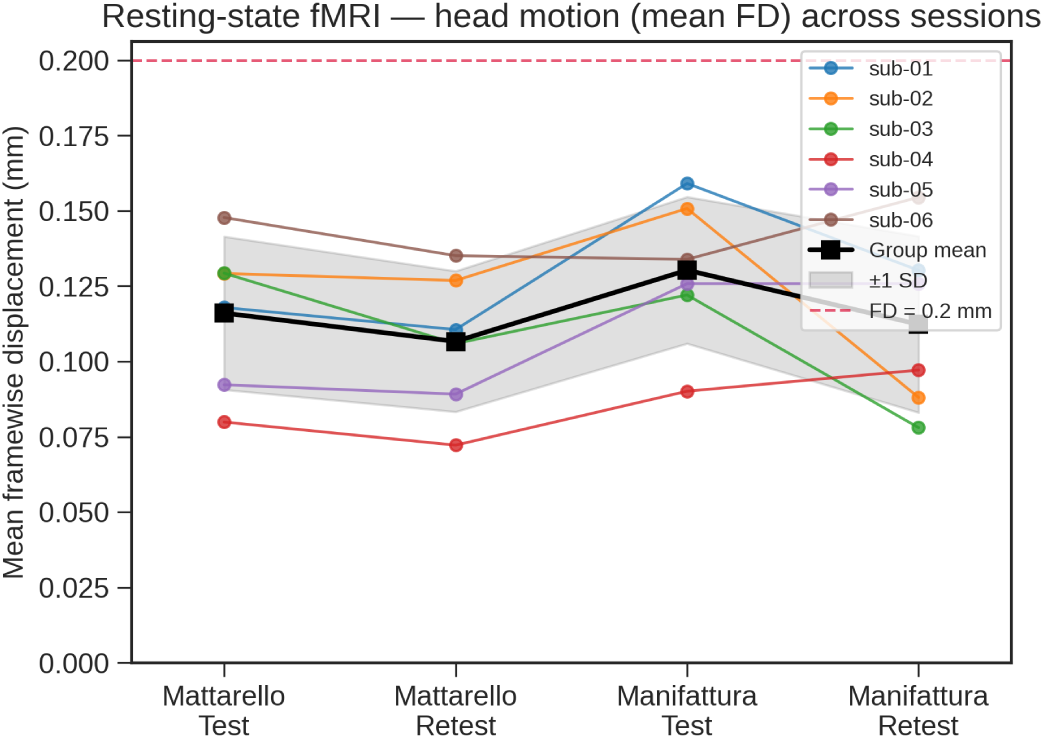
Tracking of global head motion across subjects and sessions. No significant differences were found in either of the main factors (Subject, Site).

**Fig. C2.**
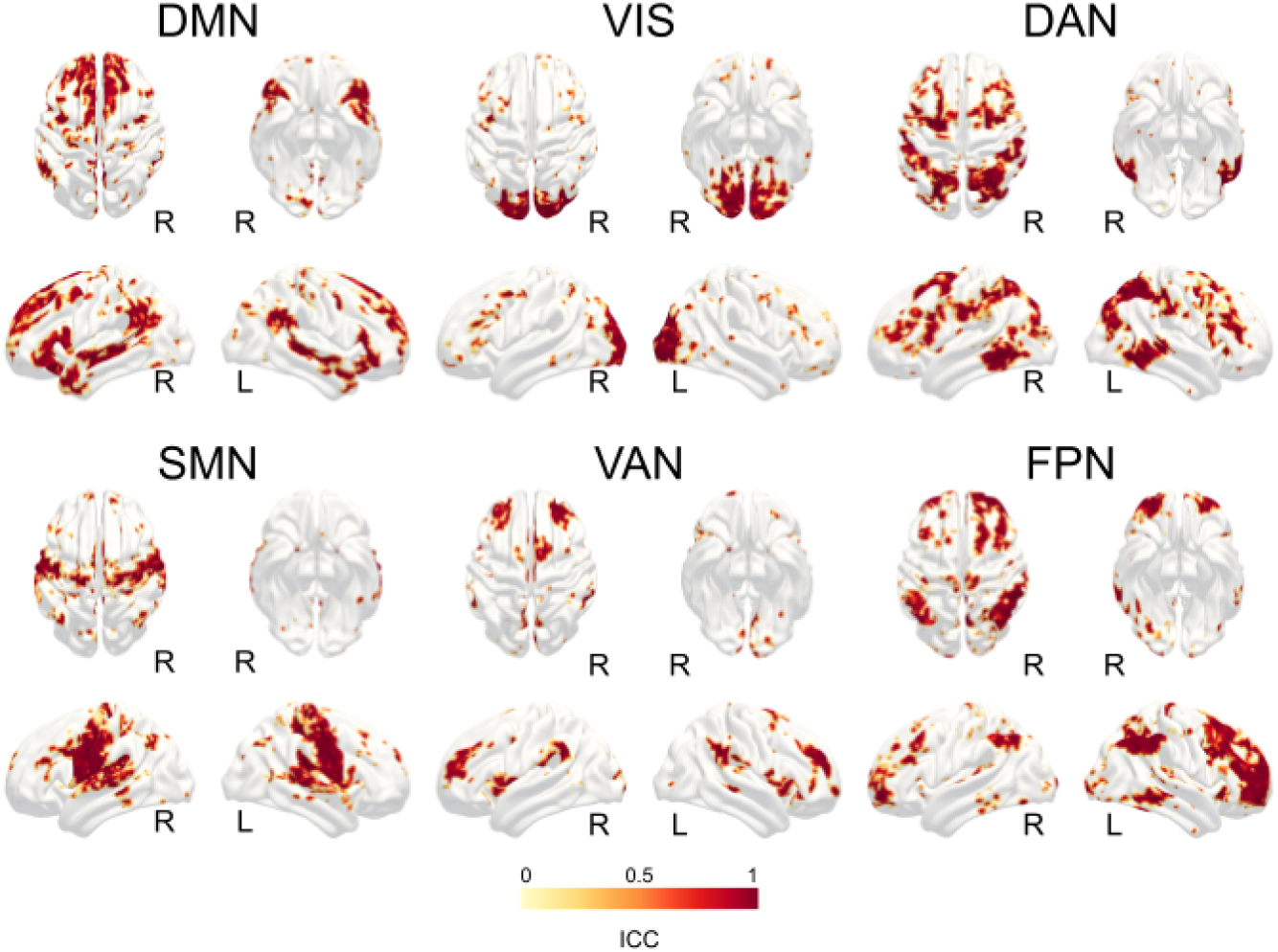
Within Site 1 reproducibility of the six major resting-state networks extracted from multiecho EPI imaging. Abbreviations: DMN, default mode network; VIS, visual network; DAN, dorsal attention network; SMN, somato sensory motor network; VAN, ventral attention network; FPN, fronto parietal network.

**Fig. C3.**
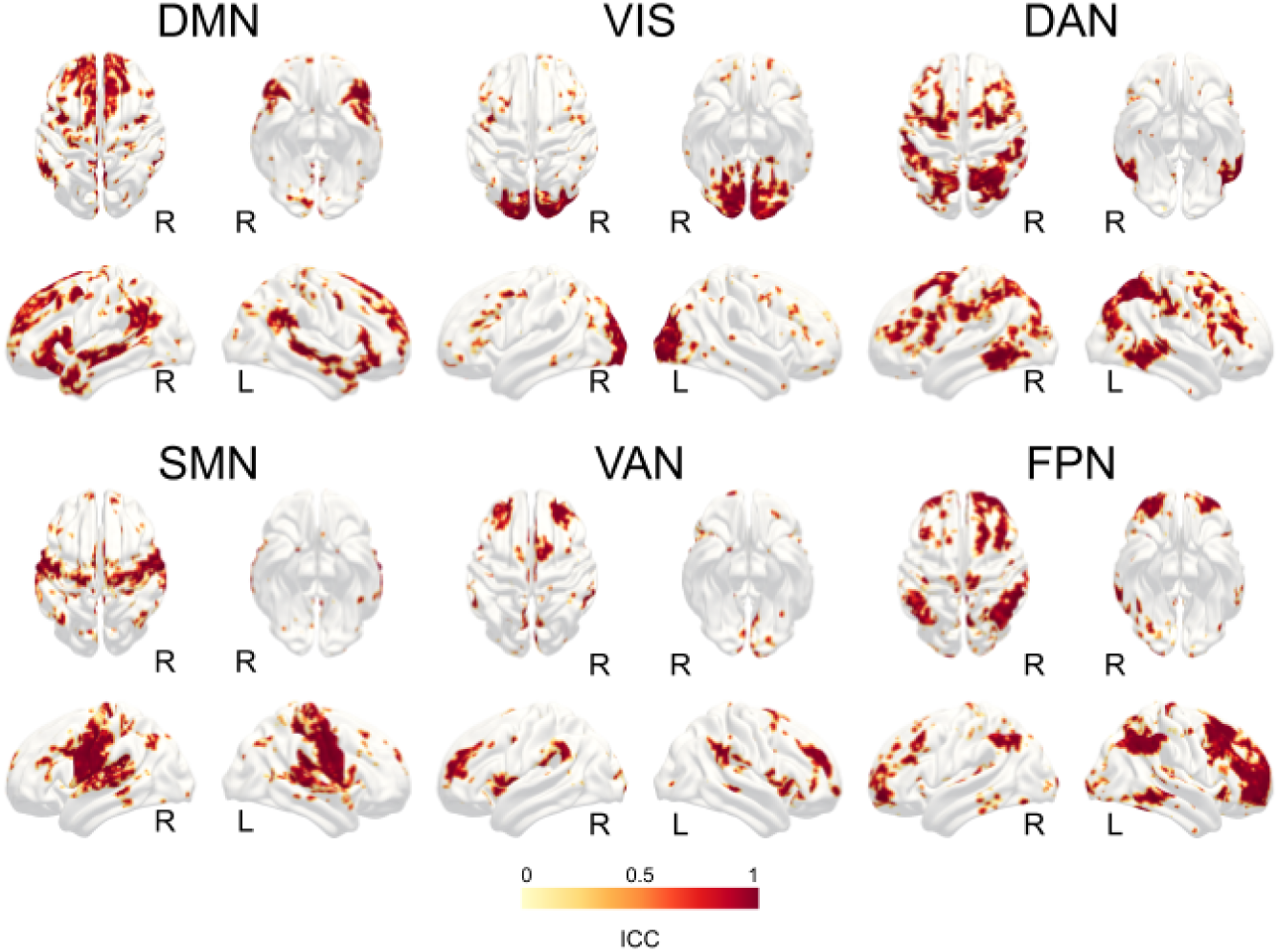
Within Site 2 reproducibility of the six major resting-state networks extracted from multiecho EPI imaging. Abbreviations: DMN, default mode network; VIS, visual network; DAN, dorsal attention network; SMN, somato sensory motor network; VAN, ventral attention network; FPN, fronto parietal network.

### Task-based fMRI

**Fig. F1.**
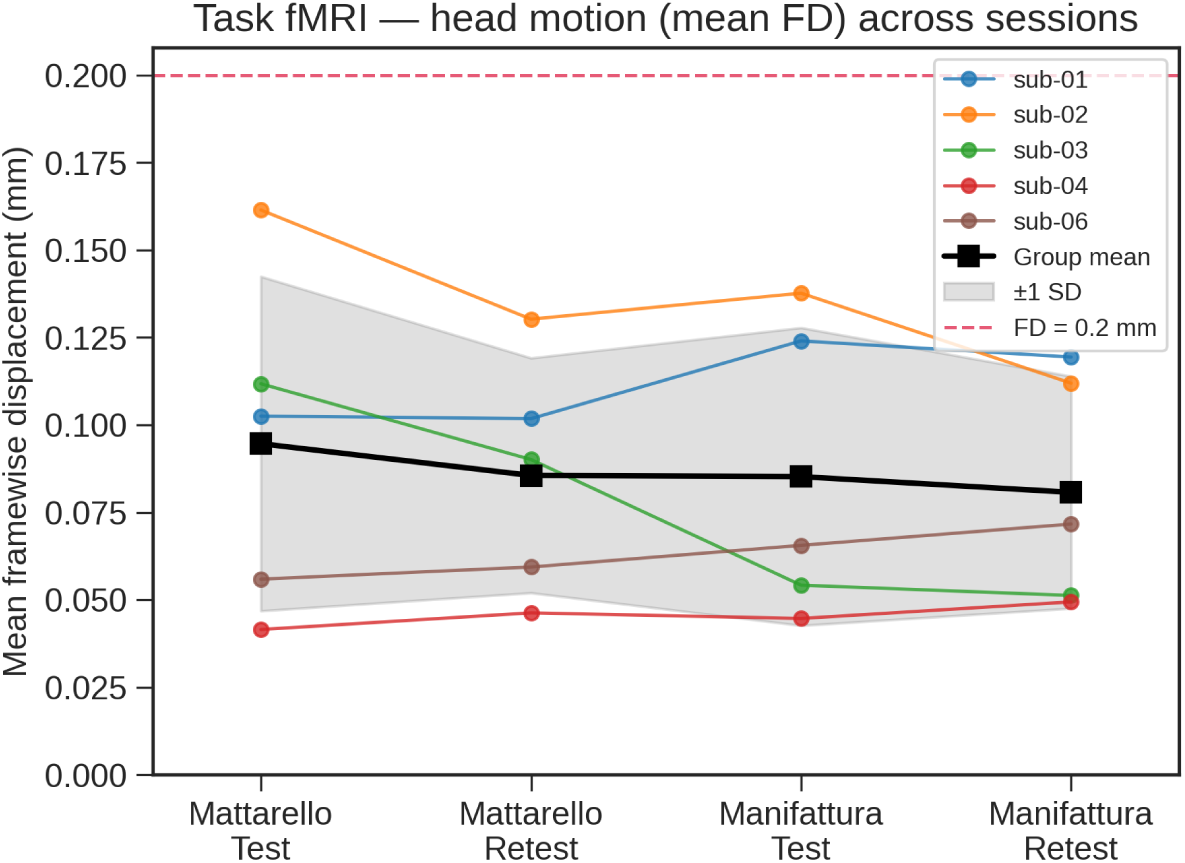
Tracking of global head motion across subjects and sessions. No significant differences were found in either of the main factors (Subject, Site).

**Fig. F2.**
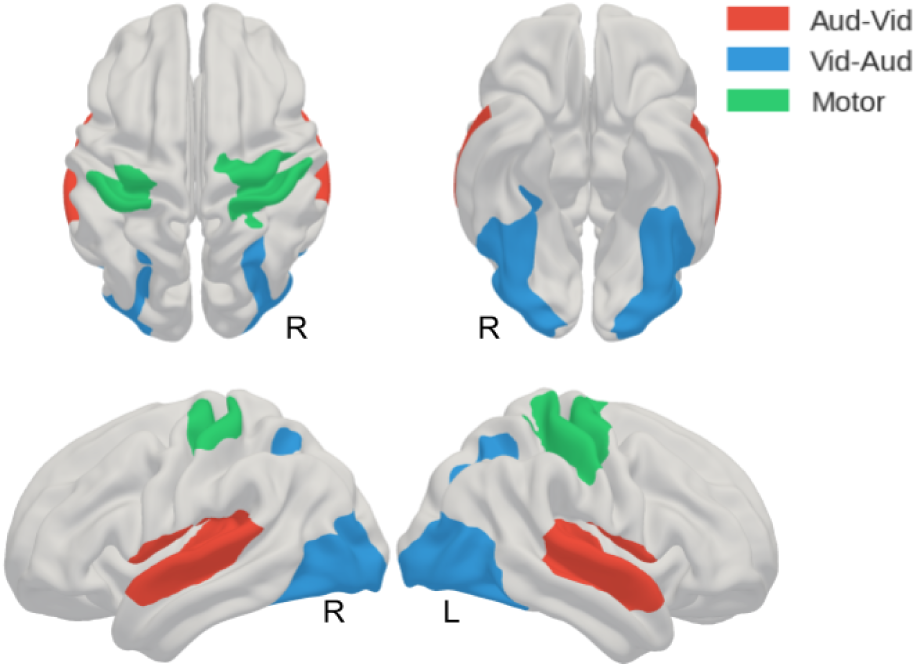
Surface rendering of the sensorimotor ROIs used to assess within and between sites reproducibility of fMRI contrasts.

**Fig. F3.**
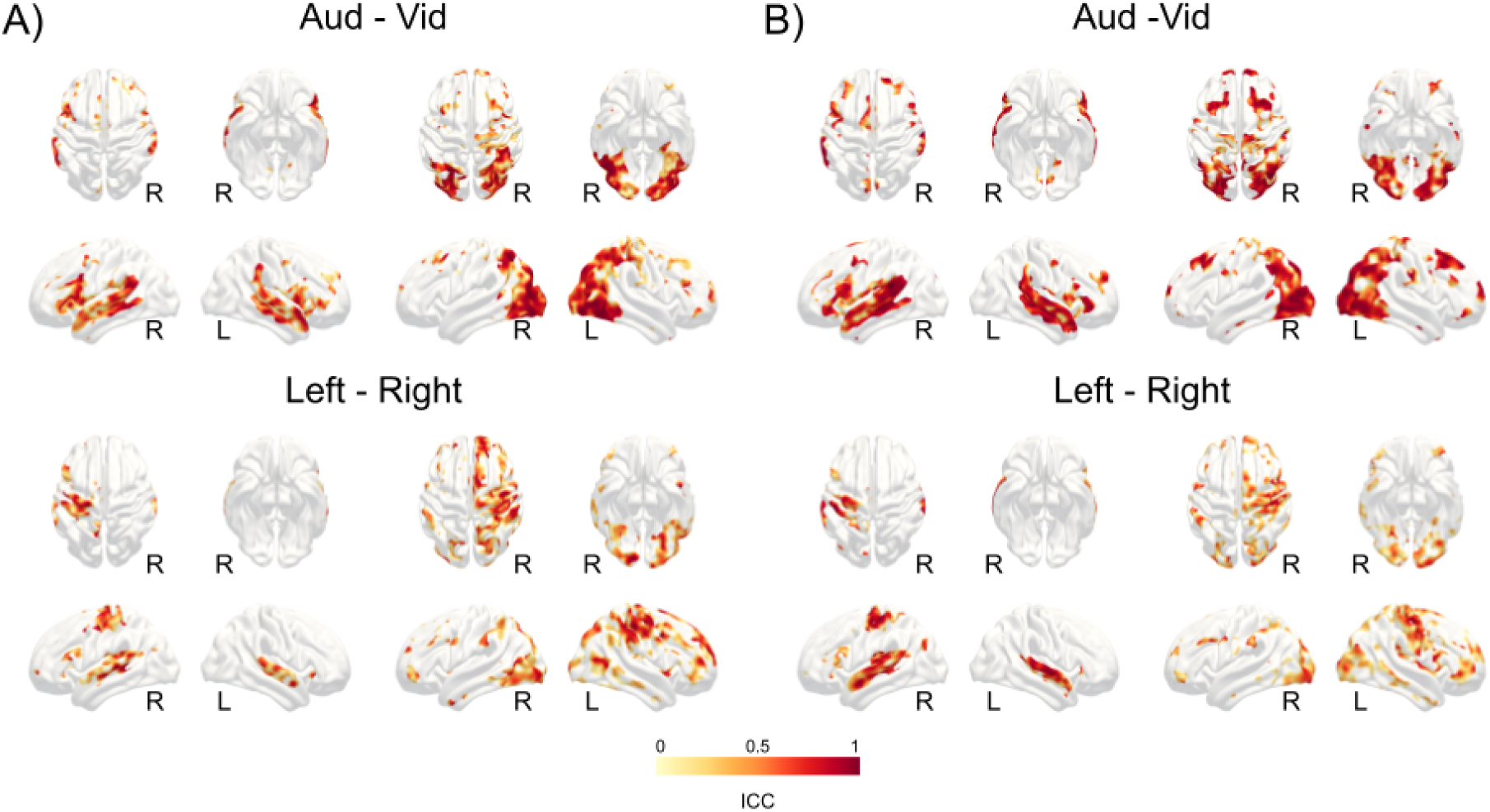
Within site reproducibility of activation strength for audio-visual and left-right contrasts. Panel A): within Site 1; panel B): within Site 2

**Tab. T1.**
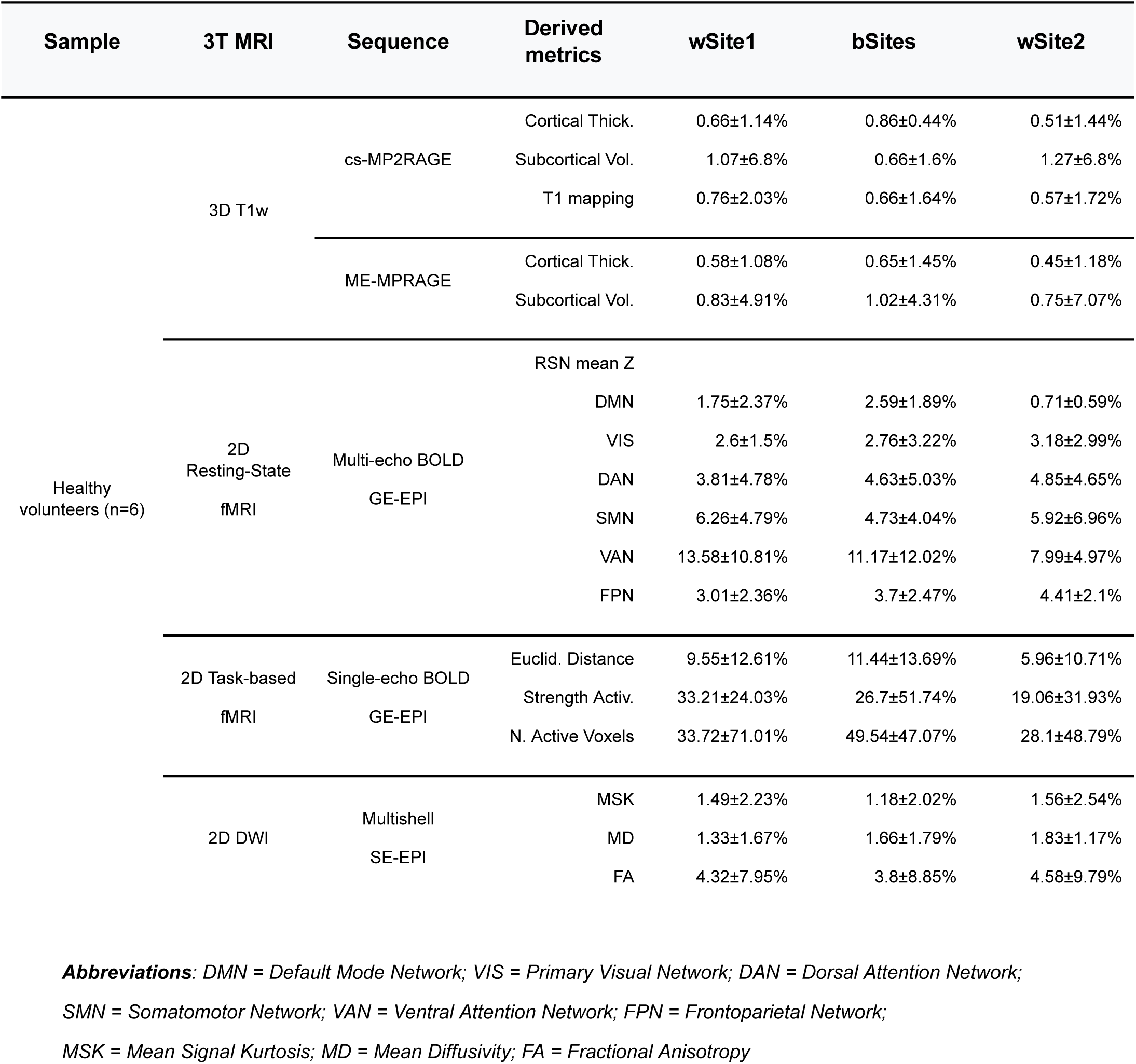
Overview of the experimental design and summary result for the three test-retest sessions.

## Notes

### Competing Interest Statement

The authors have declared no competing interest.

### Author Declarations

Ethical Committee of the University of Trento gave ethical approval for this work

